# Renin Angiotensin System Inhibition and Susceptibility and Outcomes from COVID-19: A Systematic Review and Meta-analysis of 69,200 COVID-19 Patients

**DOI:** 10.1101/2020.10.03.20206375

**Authors:** Yi Zhang, Shikai Yu, Yawei Xu, Bryan Williams

**Author notes:** authors contributed equally to this manuscript (Yi Zhang, Shikai Yu). **Corresponding authors:** Professor Yawei Xu, Cardiac center, Shanghai Tenth People’s Hospital, Tongji University, 301 Middle Yanchang Road, Shanghai, China, Professor Bryan Williams, Chair of Medicine | University College London, Maple House 1^st^ Floor Suite A, 149 Tottenham Court Road, London W1T 7DN, United Kingdom.

## Abstract

**Background:** Early observational studies suggested that the use of the renin angiotensin system (RAS) inhibitors, specifically angiotensin converting enzyme inhibitors or angiotensin receptor blockers, may increase the risk of infection with SARS-CoV-2 and adversely affect the prognosis or survival of infected patients. To explore the impact of RAS inhibitor use on the risk of SARS-CoV-2 infection and the prognosis of SARS-CoV-2 infected patients, from all published studies.

**Methods and Findings:** A systematic review and meta-analysis of the use of RAS inhibitors in relation to infection with SARS-CoV-2 and/or the severity and mortality associated with COVID-19 was conducted. English language bibliographic databases PubMed, Web of Science, OVID Embase, Scopus, MedRxiv, BioRxiv, searched from Jan 1st, 2020 to July 20th, 2020. 58 observational studies (69,200 COVID-19 patients and 3,103,335 controls) were included. There was no difference in the susceptibility to SARS-CoV-2 infection between RAS inhibitor users and non-users (unadjusted OR 1.05, 95% CI 0.90 to 1.21), (adjusted OR 0.93, 95% CI 0.85 to 1.02), (adjusted HR 1.07, 95% CI 0.87 to 1.31). There was no significant difference in the severe Covid-19 case rate between RAS inhibitor users and non-users (unadjusted OR 1.05, 95% CI 0.81 to 1.36), (adjusted OR 0.76, 95% CI 0.52 to 1.12), or in mortality due to COVID-19 between RAS inhibitor users and non-users (unadjusted OR 1.12, 95% CI 0.88 to 1.44), (adjusted OR 0.97, 95% CI 0.77 to 1.23), (adjusted HR 0.62, 95% CI 0.34 to 1.14).

**Conclusions:** In the most comprehensive analysis of all available data to date, treatment with RAS inhibitors was not associated with increased risk of infection, severity of disease, or mortality due to COVID-19. The best available evidence suggests that these treatments should not be discontinued on the basis of concern about risk associated with COVID-19.

## Introduction

Following the onset of the COVID-19 pandemic, the observation that angiotensin-converting enzyme 2 (ACE2) was the binding site through which SARS-CoV-2 gained cellular entry^1^, prompted much speculation about the safety of renin angiotensin system (RAS) inhibitors, such as angiotensin converting enzyme inhibitors (ACEi) or angiotensin receptor blocker (ARBs). This concern arose because previous studies had suggested that RAS inhibitors may lead to increased ACE2 expression on the cell surface^2^, and it was assumed that this might increase the susceptibility of patients using these medications to SARS-CoV-2 infection and when infected, increase the risk of an adverse outcome from COVID-19.

RAS inhibitors are widely used to treat patients with hypertension, heart failure, chronic kidney disease and diabetes, all of which are co-morbidities associated with more severe disease and a worse outcome from COVID-19. These concerns led to many patients at risk of severe COVID-19 infection due to these co-morbidities, to discontinue their treatment with RAS inhibitors. This prompted several professional medical and scientific societies to caution against withdrawing RAS inhibitors therapy, whilst acknowledging that more data was needed.^3^ Other commentators suggested a pragmatic solution to continue ACEi or ARBs in patients at highest short term cardiovascular risk but consider stopping these treatments in the majority of patients at lower CVD risk, if they tested positive for Covid-19.^4^

Subsequently, a series of impressive observational and case-controlled studies rapidly emerged that have provided some reassurance about the safety of RAS inhibitors in the context of COVID-19 infection. However, the studies vary in size and region and no individual study has been able to address all of the key questions about the potential impact of RAS inhibitors on the susceptibility to SARS-CoV-2 infection, the severity of infection and risk of mortality. Moreover, questions have remained about the impact of hypertension *per se* on the risk of COVID-19 and the impact of RAS inhibitors in this large cohort of patients. We therefore undertook a systematic review and meta-analysis of all existing clinical data to provide the most definitive report to date on the association between RAS inhibitors and the susceptibility to SARS-CoV-2 infection (18 studies), the severity of the resulting disease (29 studies) and mortality (30 studies) due to COVID-19.

## Methods

This meta-analysis was conducted in accordance with the Preferred Reporting Items for Systematic Reviews and Meta-analyses Protocols (PRISMA) ^5^ and the Meta-analysis of Observational Studies in Epidemiology guidelines (MOOSE).^6^

### Eligibility criteria, search strategy and study selection

We performed a systematic literature search for studies of any design and in any setting in electronic bibliographic databases including PubMed, Web of Science, OVID Embase, and Scopus from Jan 1^st^, 2020 to July 20^th^, 2020, using a comprehensive search strategy as described in eAppendix 1 in the Supplement. We also manually searched preprint platforms (MedRxiv and BioRxiv), coronavirus resource centers of N Engl J Med, JAMA, BMJ and the Lancet, and checked reference lists of eligible studies to find additional studies. We limited our search by language (English). Since COVID-19 topic was a novel disease, any study that reported the associations of the susceptibility to infection with SARS-CoV-2 and severity and mortality of COVID-19 with the use of RAS inhibitors was eligible. Two authors (S.Y. and Y.Z.) independently screened and examined eligible studies. Disagreements were resolved by consensus.

### Outcomes

There were three outcomes of interest; (i) susceptibility to infection with SARS-Cov-2; (ii) the severity of COVID-19 infection; and (iii) mortality from COVID-19. Cases of COVID-19 were defined as those with a positive test result from a real-time reverse-transcriptase-polymerase-chain-reaction (RT-PCR) assay of nasopharyngeal swab samples. Definitions of disease severity of COVID-19 varied among studies (eTable 4 in the Supplement) and it was not feasible to provide a single consistent definition. Most studies from China used the National Health Commission of China guideline, which categorized patients into four types of severity (mild, common, severe, and critical). In these Chinese studies, severe and critical were classified as severe cases and mild and common as non-severe cases. The other studies defined severe cases as admission to an ICU and/or death and/or long hospitalization (eTable 4 in the Supplement). We pooled the result across all included studies based on different definitions of severity and performed subgroup analyses according to the countries of origin (China and Non-China).

### Data extraction and risk of bias

Two reviewers (S.Y. and Y.Z.) independently extracted study data and evaluated studies for risk of bias. The following information was extracted from each study: study identifier, study design, country, study size, mean/median age, male sex proportion, quantitative outcomes, effect estimators (Odds ratios [ORs] or Hazard ratios [HRs]) and adjusted covariates if available. The comparison of outcomes comparing ACEi or ARB treatment with non-ACEi or ARB treatment was our primary interest, but several included studies reported data for ACEi and ARB separately. For those studies, we used the additional data for ACEi and ARB to calculate the corresponding data for ACEi or ARB as a single entity, which was a reasonable approach considering the very small possibility of combined use of ACEi and ARB in clinical practice.

Newcastle-Ottawa Scale (NOS)^7^ was used to evaluate the risk of bias (or quality) of each observational study. NOS was calculated according to three major components: selection (0-4 points), comparability (0-2 points) and outcome (0-3 points). A total score of 7 or more was considered as low risk of bias. We planned to use the Cochrane Risk of Bias tool to evaluate the risk of bias in any randomized clinical trials (RCTs), but only one retrospective interim analysis of an unfinished RCT study was identified for this meta-analysis which was treated as a cohort study and evaluated with NOS.

### Statistical analysis

The primary analysis estimated the associations of susceptibility to infection with SARS-Cov-2 and the severity and mortality from COVID-19 with the use of ACEi or ARB. Pooled unadjusted ORs were calculated with the Mantel-Haenszel method with a random-effects or fixed-effects model (wherever appropriate), pooled adjusted ORs were calculated with the generic inverse variance method with a random-effects or fixed-effects model. In terms of mortality, some studies reported ORs, others reported HRs, thus we pooled ORs and HRs separately. We performed a series of subgroup analyses in hypertensive patients, Chinese/non-Chinese, case-controlled studies, and non-severe and severe patients, wherever data was available for the three outcomes of interest. Additionally, we calculated the pooled estimates for outcomes with ACEi and ARB separately.

Inter-study heterogeneity was assessed with the χ^2^ based-Q-statistic test and I^2^ statistic. I^2^ < 25%, 25%-50% and >50% suggested low, moderate, and high heterogeneity, respectively. We examined the potential publication bias using Funnel plots and Egger’s test. Leave-one-out sensitivity analyses were applied to evaluate the robustness of our findings after excluding each included study on the pooled results. All statistical analyses were conducted using Reviewer Manager 5.3 (The Nordic Cochrane Center, The Cochrane Collaboration, 2018, Copenhagen, Denmark) and STATA version 12.0 (Stata Corp., College Station, TX).

### Patients and Public Involvement

This was a systematic review and meta-analysis of previously published data and there was no involvement of patients or the public in this study.

## Results

The search strategy identified 536 published articles from the PubMed, 600 from the Web of Science, 424 from the OVID Embase and 390 from the Scopus, and 24 electronically published articles from the preprint platforms (Figure 1). After duplication and eligibility screening, 81 articles remained. 23 articles were further omitted because of unrelated topic, unavailable or overlapping data, ineligible design, or journal retraction. A total of 58 studies were included in the final meta-analysis, 18 provided susceptibility data (7 with adjusted ORs and 3 with adjusted HRs), 29 provided disease severity data (13 with adjusted OR and 1 with adjusted HRs), and 30 provided mortality data (8 with adjusted ORs and 3 with adjusted HRs).

**Figure 1.**
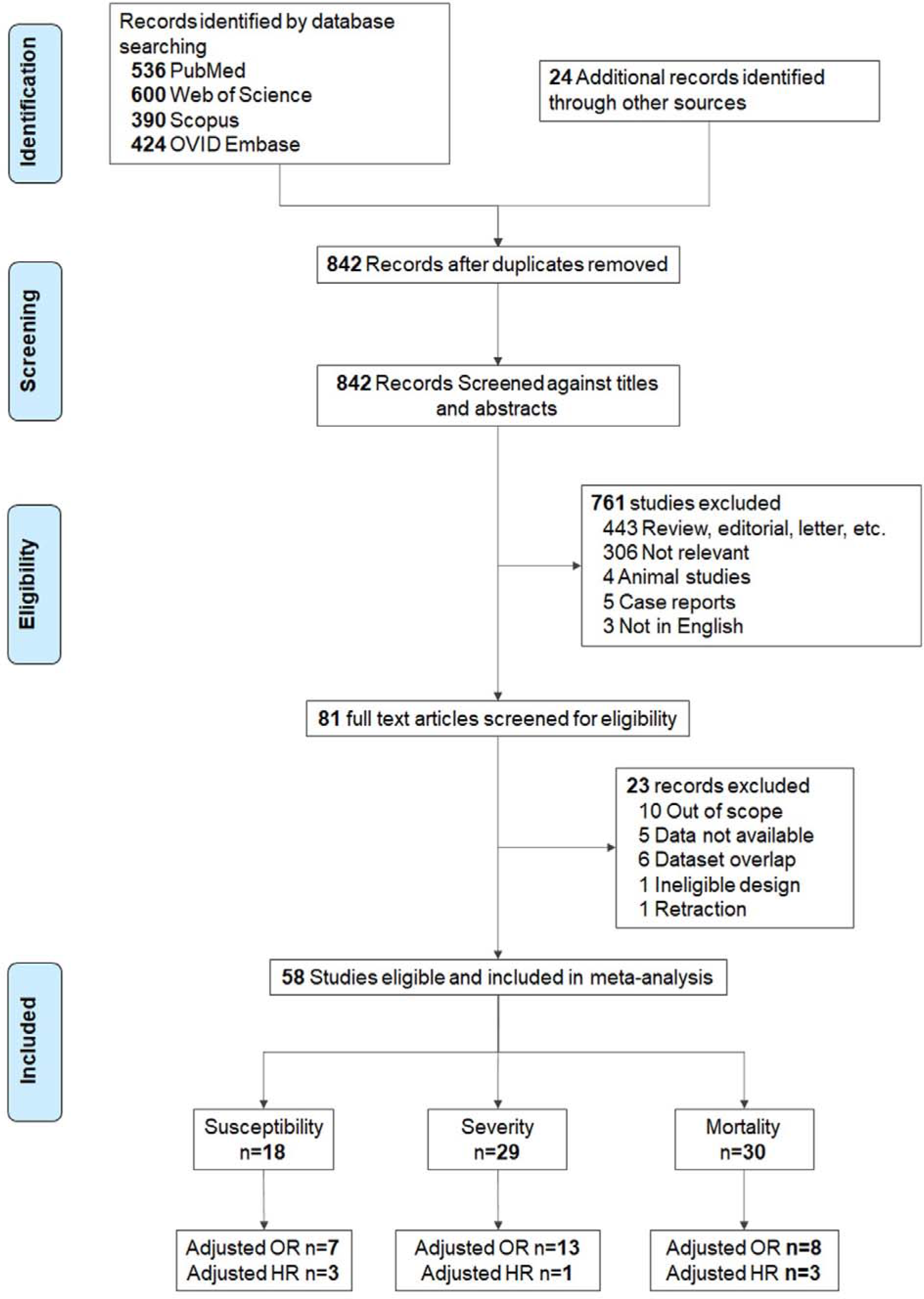
Flowchart of study selection

The Table describes the cohort characteristics of the included studies. Our analysis comprised 69,200 COVID-19 patients and 3,103,335 controls from 58 studies (35 published in journals and 23 available as preprints via MedRxiv). Since not all studies provided three major outcomes of interest i.e. susceptibility to infection, disease severity and mortality, we collated the studies into 3 groups, according to the availability of data for the three major outcomes (Table).

**Table.**
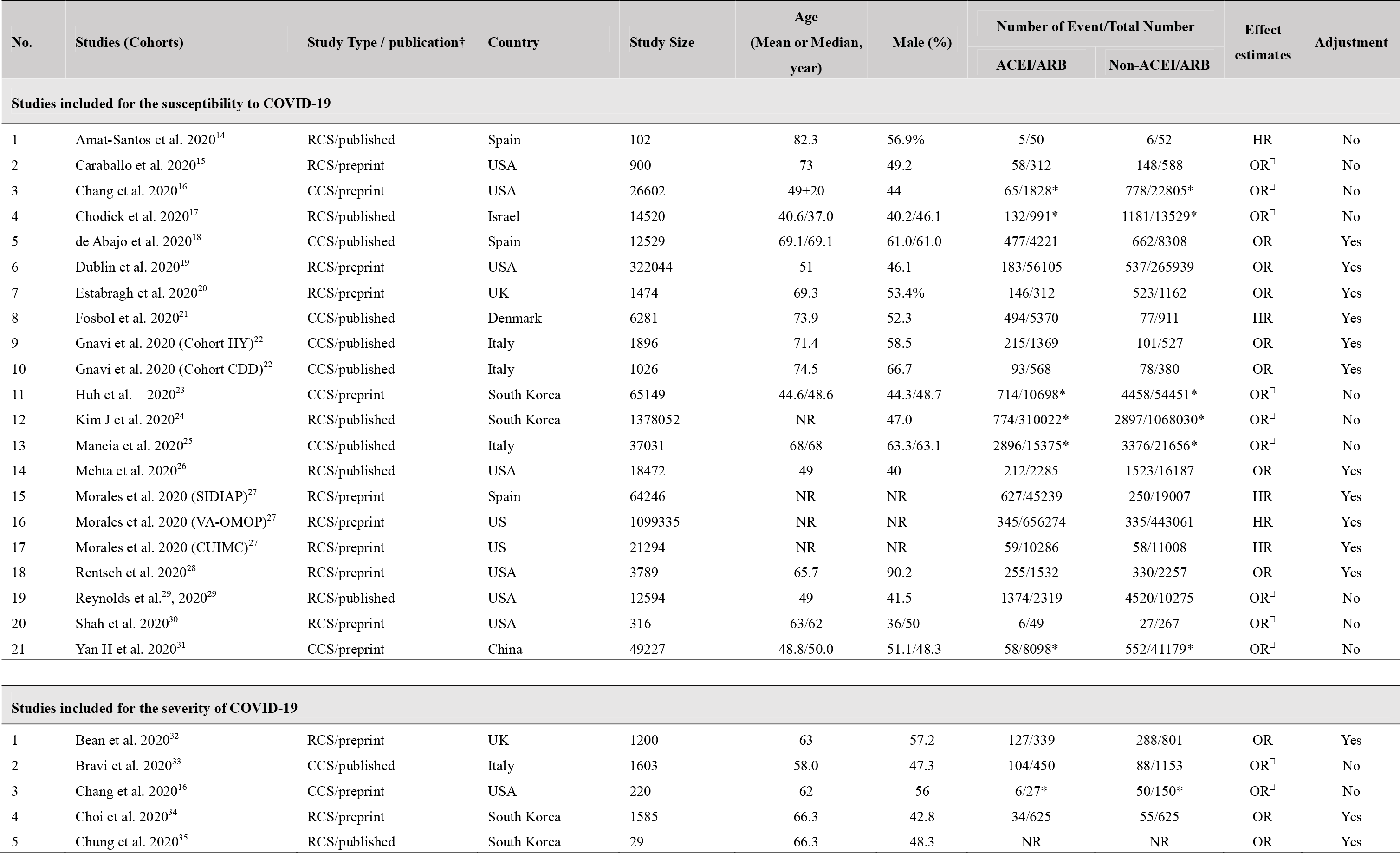
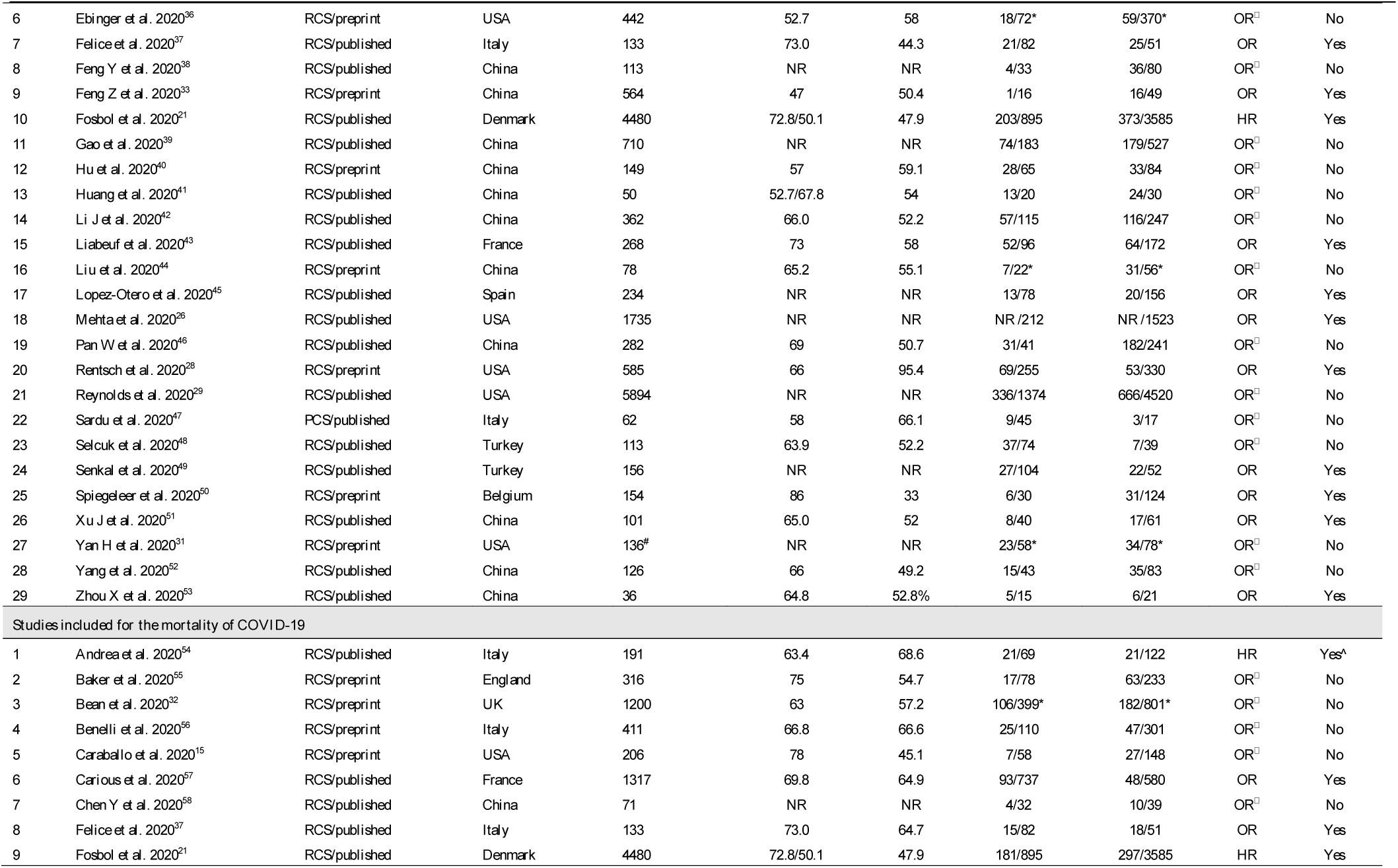
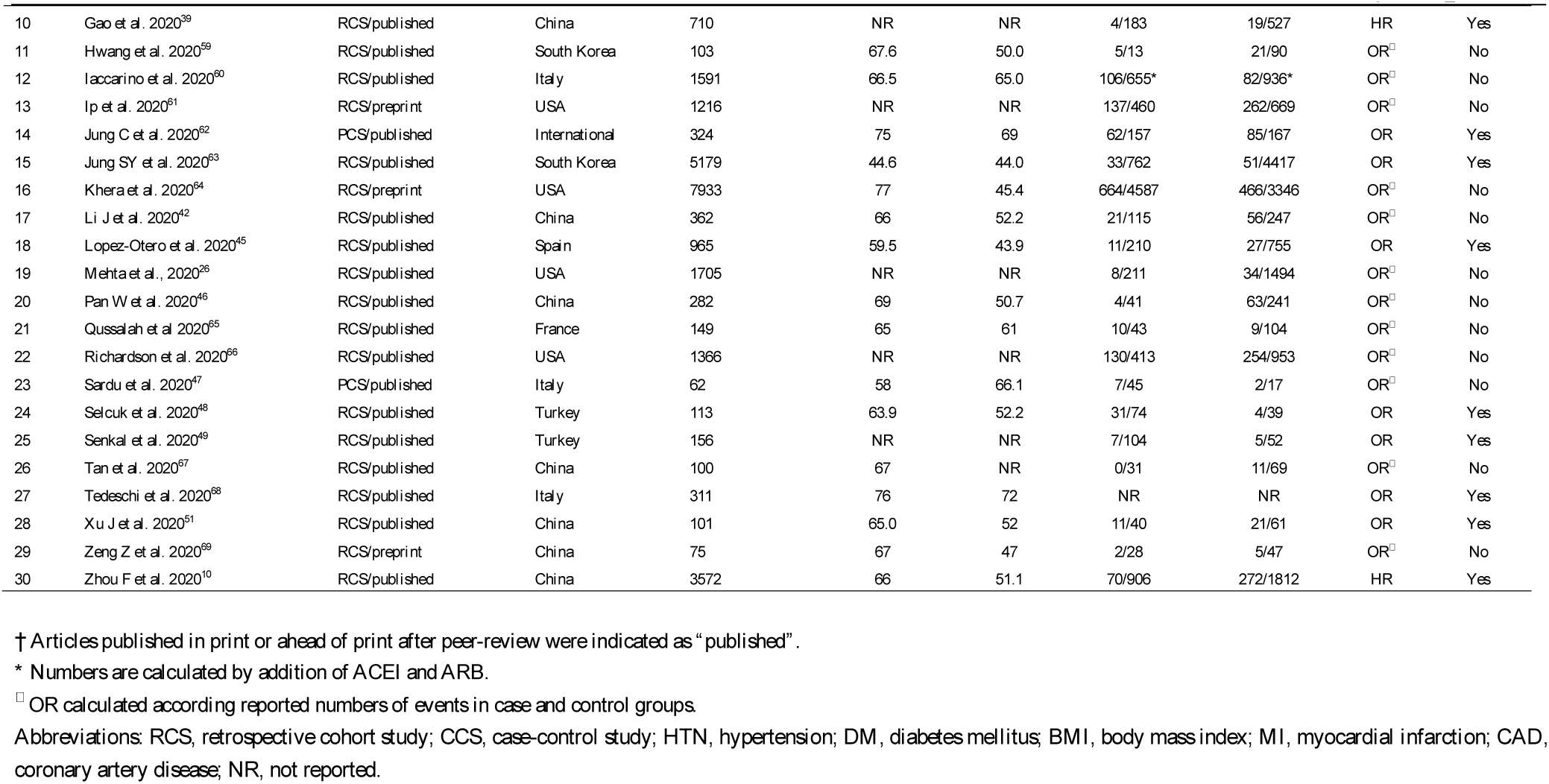
Main characteristics of studies included in these analyses

**Table. Main characteristics of studies included in these analyses**
| No. | Studies (Cohorts) | Study Type / publication† | Country | Study Size | Age<br>(Mean or Median,<br>year) | Male (%) | Number of Event/Total Number |  | Effect<br>estimates | Adjustment |
| --- | --- | --- | --- | --- | --- | --- | --- | --- | --- | --- |
|  |  |  |  |  |  |  | ACEI/ARB | Non-ACEI/ARB |  |  |
| Studies included for the susceptibility to COVID-19 |  |  |  |  |  |  |  |  |  |  |
| 1 | Amat-Santos et al. 2020<sup>14</sup> | RCS/published | Spain | 102 | 82.3 | 56.9% | 5/50 | 6/52 | HR | No |
| 2 | Caraballo et al. 2020<sup>15</sup> | RCS/preprint | USA | 900 | 73 | 49.2 | 58/312 | 148/588 | OR<sup>□</sup> | No |
| 3 | Chang et al. 2020<sup>16</sup> | CCS/preprint | USA | 26602 | 49±20 | 44 | 65/1828* | 778/22805* | OR<sup>□</sup> | No |
| 4 | Chodick et al. 2020<sup>17</sup> | RCS/published | Israel | 14520 | 40.6/37.0 | 40.2/46.1 | 132/991* | 1181/13529* | OR<sup>□</sup> | No |
| 5 | de Abajo et al. 2020<sup>18</sup> | CCS/published | Spain | 12529 | 69.1/69.1 | 61.0/61.0 | 477/4221 | 662/8308 | OR | Yes |
| 6 | Dublin et al. 2020<sup>19</sup> | RCS/preprint | USA | 322044 | 51 | 46.1 | 183/56105 | 537/265939 | OR | Yes |
| 7 | Estabragh et al. 2020<sup>20</sup> | RCS/preprint | UK | 1474 | 69.3 | 53.4% | 146/312 | 523/1162 | OR | Yes |
| 8 | Fosbol et al. 2020<sup>21</sup> | CCS/published | Denmark | 6281 | 73.9 | 52.3 | 494/5370 | 77/911 | HR | Yes |
| 9 | Gnavi et al. 2020 (Cohort HY)<sup>22</sup> | CCS/published | Italy | 1896 | 71.4 | 58.5 | 215/1369 | 101/527 | OR | Yes |
| 10 | Gnavi et al. 2020 (Cohort CDD)<sup>22</sup> | CCS/published | Italy | 1026 | 74.5 | 66.7 | 93/568 | 78/380 | OR | Yes |
| 11 | Huh et al. 2020<sup>23</sup> | CCS/preprint | South Korea | 65149 | 44.6/48.6 | 44.3/48.7 | 714/10698* | 4458/54451* | OR<sup>□</sup> | No |
| 12 | Kim J et al. 2020<sup>24</sup> | RCS/published | South Korea | 1378052 | NR | 47.0 | 774/310022* | 2897/1068030* | OR<sup>□</sup> | No |
| 13 | Mancia et al. 2020<sup>25</sup> | CCS/published | Italy | 37031 | 68/68 | 63.3/63.1 | 2896/15375* | 3376/21656* | OR<sup>□</sup> | No |
| 14 | Mehta et al. 2020<sup>26</sup> | RCS/published | USA | 18472 | 49 | 40 | 212/2285 | 1523/16187 | OR | Yes |
| 15 | Morales et al. 2020 (SIDIAP)<sup>27</sup> | RCS/preprint | Spain | 64246 | NR | NR | 627/45239 | 250/19007 | HR | Yes |
| 16 | Morales et al. 2020 (VA-OMOP)<sup>27</sup> | RCS/preprint | US | 1099335 | NR | NR | 345/656274 | 335/443061 | HR | Yes |
| 17 | Morales et al. 2020 (CUIMC)<sup>27</sup> | RCS/preprint | US | 21294 | NR | NR | 59/10286 | 58/11008 | HR | Yes |
| 18 | Rentsch et al. 2020<sup>28</sup> | RCS/preprint | USA | 3789 | 65.7 | 90.2 | 255/1532 | 330/2257 | OR | Yes |
| 19 | Reynolds et al.<sup>29</sup>, 2020<sup>29</sup> | RCS/published | USA | 12594 | 49 | 41.5 | 1374/2319 | 4520/10275 | OR<sup>□</sup> | No |
| 20 | Shah et al. 2020<sup>30</sup> | RCS/preprint | USA | 316 | 63/62 | 36/50 | 6/49 | 27/267 | OR<sup>□</sup> | No |
| 21 | Yan H et al. 2020<sup>31</sup> | CCS/preprint | China | 49227 | 48.8/50.0 | 51.1/48.3 | 58/8098* | 552/41179* | OR<sup>□</sup> | No |
| **Studies included for the severity of COVID-19** |  |  |  |  |  |  |  |  |  |  |
| 1 | Bean et al. 2020<sup>32</sup> | RCS/preprint | UK | 1200 | 63 | 57.2 | 127/339 | 288/801 | OR | Yes |
| 2 | Bravi et al. 2020<sup>33</sup> | CCS/published | Italy | 1603 | 58.0 | 47.3 | 104/450 | 88/1153 | OR<sup>□</sup> | No |
| 3 | Chang et al. 2020<sup>16</sup> | CCS/preprint | USA | 220 | 62 | 56 | 6/27* | 50/150* | OR<sup>□</sup> | No |
| 4 | Choi et al. 2020<sup>34</sup> | RCS/preprint | South Korea | 1585 | 66.3 | 42.8 | 34/625 | 55/625 | OR | Yes |
| 5 | Chung et al. 2020<sup>35</sup> | RCS/published | South Korea | 29 | 66.3 | 48.3 | NR | NR | OR | Yes |
| 6 | Ebinger et al. 2020<sup>36</sup> | RCS/preprint | USA | 442 | 52.7 | 58 | 18/72* | 59/370* | OR<sup>□</sup> | No |
| 7 | Felice et al. 2020<sup>37</sup> | RCS/published | Italy | 133 | 73.0 | 44.3 | 21/82 | 25/51 | OR | Yes |
| 8 | Feng Y et al. 2020<sup>38</sup> | RCS/published | China | 113 | NR | NR | 4/33 | 36/80 | OR<sup>□</sup> | No |
| 9 | Feng Z et al. 2020<sup>33</sup> | RCS/preprint | China | 564 | 47 | 50.4 | 1/16 | 16/49 | OR | Yes |
| 10 | Fosbol et al. 2020<sup>21</sup> | RCS/published | Denmark | 4480 | 72.8/50.1 | 47.9 | 203/895 | 373/3585 | HR | Yes |
| 11 | Gao et al. 2020<sup>39</sup> | RCS/published | China | 710 | NR | NR | 74/183 | 179/527 | OR<sup>□</sup> | No |
| 12 | Hu et al. 2020<sup>40</sup> | RCS/preprint | China | 149 | 57 | 59.1 | 28/65 | 33/84 | OR<sup>□</sup> | No |
| 13 | Huang et al. 2020<sup>41</sup> | RCS/published | China | 50 | 52.7/67.8 | 54 | 13/20 | 24/30 | OR<sup>□</sup> | No |
| 14 | Li J et al. 2020<sup>42</sup> | RCS/published | China | 362 | 66.0 | 52.2 | 57/115 | 116/247 | OR<sup>□</sup> | No |
| 15 | Liabeuf et al. 2020<sup>43</sup> | RCS/published | France | 268 | 73 | 58 | 52/96 | 64/172 | OR | Yes |
| 16 | Liu et al. 2020<sup>44</sup> | RCS/preprint | China | 78 | 65.2 | 55.1 | 7/22* | 31/56* | OR<sup>□</sup> | No |
| 17 | Lopez-Otero et al. 2020<sup>45</sup> | RCS/published | Spain | 234 | NR | NR | 13/78 | 20/156 | OR | Yes |
| 18 | Mehta et al. 2020<sup>26</sup> | RCS/published | USA | 1735 | NR | NR | NR /212 | NR /1523 | OR | Yes |
| 19 | Pan W et al. 2020<sup>46</sup> | RCS/published | China | 282 | 69 | 50.7 | 31/41 | 182/241 | OR<sup>□</sup> | No |
| 20 | Rentsch et al. 2020<sup>28</sup> | RCS/preprint | USA | 585 | 66 | 95.4 | 69/255 | 53/330 | OR | Yes |
| 21 | Reynolds et al. 2020<sup>29</sup> | RCS/published | USA | 5894 | NR | NR | 336/1374 | 666/4520 | OR<sup>□</sup> | No |
| 22 | Sardu et al. 2020<sup>47</sup> | PCS/published | Italy | 62 | 58 | 66.1 | 9/45 | 3/17 | OR<sup>□</sup> | No |
| 23 | Selcuk et al. 2020<sup>48</sup> | RCS/published | Turkey | 113 | 63.9 | 52.2 | 37/74 | 7/39 | OR<sup>□</sup> | No |
| 24 | Senkal et al. 2020<sup>49</sup> | RCS/published | Turkey | 156 | NR | NR | 27/104 | 22/52 | OR | Yes |
| 25 | Spiegeleer et al. 2020<sup>50</sup> | RCS/preprint | Belgium | 154 | 86 | 33 | 6/30 | 31/124 | OR | Yes |
| 26 | Xu J et al. 2020<sup>51</sup> | RCS/published | China | 101 | 65.0 | 52 | 8/40 | 17/61 | OR | Yes |
| 27 | Yan H et al. 2020<sup>31</sup> | RCS/preprint | USA | 136<sup>#</sup> | NR | NR | 23/58* | 34/78* | OR<sup>□</sup> | No |
| 28 | Yang et al. 2020<sup>52</sup> | RCS/published | China | 126 | 66 | 49.2 | 15/43 | 35/83 | OR<sup>□</sup> | No |
| 29 | Zhou X et al. 2020<sup>53</sup> | RCS/published | China | 36 | 64.8 | 52.8% | 5/15 | 6/21 | OR | Yes |
| **Studies included for the mortality of COVID-19** |  |  |  |  |  |  |  |  |  |  |
| 1 | Andrea et al. 2020<sup>54</sup> | RCS/published | Italy | 191 | 63.4 | 68.6 | 21/69 | 21/122 | HR | Yes^ |
| 2 | Baker et al. 2020<sup>55</sup> | RCS/preprint | England | 316 | 75 | 54.7 | 17/78 | 63/233 | OR<sup>□</sup> | No |
| 3 | Bean et al. 2020<sup>32</sup> | RCS/preprint | UK | 1200 | 63 | 57.2 | 106/399* | 182/801* | OR<sup>□</sup> | No |
| 4 | Benelli et al. 2020<sup>56</sup> | RCS/preprint | Italy | 411 | 66.8 | 66.6 | 25/110 | 47/301 | OR<sup>□</sup> | No |
| 5 | Caraballo et al. 2020<sup>15</sup> | RCS/preprint | USA | 206 | 78 | 45.1 | 7/58 | 27/148 | OR<sup>□</sup> | No |
| 6 | Cariou et al. 2020<sup>57</sup> | RCS/published | France | 1317 | 69.8 | 64.9 | 93/737 | 48/580 | OR | Yes |
| 7 | Chen Y et al. 2020<sup>58</sup> | RCS/published | China | 71 | NR | NR | 4/32 | 10/39 | OR<sup>□</sup> | No |
| 8 | Felice et al. 2020<sup>37</sup> | RCS/published | Italy | 133 | 73.0 | 64.7 | 15/82 | 18/51 | OR | Yes |
| 9 | Fosbol et al. 2020<sup>21</sup> | RCS/published | Denmark | 4480 | 72.8/50.1 | 47.9 | 181/895 | 297/3585 | HR | Yes |
| 10 | Gao et al. 2020<sup>39</sup> | RCS/published | China | 710 | NR | NR | 4/183 | 19/527 | HR | Yes |
| 11 | Hwang et al. 2020<sup>59</sup> | RCS/published | South Korea | 103 | 67.6 | 50.0 | 5/13 | 21/90 | OR<sup>□</sup> | No |
| 12 | Iaccarino et al. 2020<sup>60</sup> | RCS/published | Italy | 1591 | 66.5 | 65.0 | 106/655* | 82/936* | OR<sup>□</sup> | No |
| 13 | Ip et al. 2020<sup>61</sup> | RCS/preprint | USA | 1216 | NR | NR | 137/460 | 262/669 | OR<sup>□</sup> | No |
| 14 | Jung C et al. 2020<sup>62</sup> | PCS/published | International | 324 | 75 | 69 | 62/157 | 85/167 | OR | Yes |
| 15 | Jung SY et al. 2020<sup>63</sup> | RCS/published | South Korea | 5179 | 44.6 | 44.0 | 33/762 | 51/4417 | OR | Yes |
| 16 | Khera et al. 2020<sup>64</sup> | RCS/preprint | USA | 7933 | 77 | 45.4 | 664/4587 | 466/3346 | OR<sup>□</sup> | No |
| 17 | Li J et al. 2020<sup>42</sup> | RCS/published | China | 362 | 66 | 52.2 | 21/115 | 56/247 | OR<sup>□</sup> | No |
| 18 | Lopez-Otero et al. 2020<sup>45</sup> | RCS/published | Spain | 965 | 59.5 | 43.9 | 11/210 | 27/755 | OR | Yes |
| 19 | Mehta et al., 2020<sup>26</sup> | RCS/published | USA | 1705 | NR | NR | 8/211 | 34/1494 | OR<sup>□</sup> | No |
| 20 | Pan W et al. 2020<sup>46</sup> | RCS/published | China | 282 | 69 | 50.7 | 4/41 | 63/241 | OR<sup>□</sup> | No |
| 21 | Qussalah et al 2020<sup>65</sup> | RCS/published | France | 149 | 65 | 61 | 10/43 | 9/104 | OR<sup>□</sup> | No |
| 22 | Richardson et al. 2020<sup>66</sup> | RCS/published | USA | 1366 | NR | NR | 130/413 | 254/953 | OR<sup>□</sup> | No |
| 23 | Sardu et al. 2020<sup>47</sup> | PCS/published | Italy | 62 | 58 | 66.1 | 7/45 | 2/17 | OR<sup>□</sup> | No |
| 24 | Selcuk et al. 2020<sup>48</sup> | RCS/published | Turkey | 113 | 63.9 | 52.2 | 31/74 | 4/39 | OR | Yes |
| 25 | Senkal et al. 2020<sup>49</sup> | RCS/published | Turkey | 156 | NR | NR | 7/104 | 5/52 | OR | Yes |
| 26 | Tan et al. 2020<sup>67</sup> | RCS/published | China | 100 | 67 | NR | 0/31 | 11/69 | OR<sup>□</sup> | No |
| 27 | Tedeschi et al. 2020<sup>68</sup> | RCS/published | Italy | 311 | 76 | 72 | NR | NR | OR | Yes |
| 28 | Xu J et al. 2020<sup>51</sup> | RCS/published | China | 101 | 65.0 | 52 | 11/40 | 21/61 | OR | Yes |
| 29 | Zeng Z et al. 2020<sup>69</sup> | RCS/preprint | China | 75 | 67 | 47 | 2/28 | 5/47 | OR<sup>□</sup> | No |
| 30 | Zhou F et al. 2020<sup>10</sup> | RCS/published | China | 3572 | 66 | 51.1 | 70/906 | 272/1812 | HR | Yes |

† Articles published in print or ahead of print after peer-review were indicated as “published”.
\* Numbers are calculated by addition of ACEI and ARB.
□ OR calculated according reported numbers of events in case and control groups.
Abbreviations: RCS, retrospective cohort study; CCS, case-control study; HTN, hypertension; DM, diabetes mellitus; BMI, body mass index; MI, myocardial infarction; CAD, coronary artery disease; NR, not reported.

### Use of RAS inhibitors and susceptibility to infection with SARS-Cov-2

Eighteen studies (3,128,679 participants) were included in the susceptibility analysis, of which 7 reported adjusted ORs and 3 reported adjusted HRs. The adjustment strategies for individual studies are listed in the Table. There was no significant difference in the incidence of COVID-19 infection between RAS inhibitor users and non-users, before and after adjustment (unadjusted OR 1.05, 0.90 to 1.21; adjusted OR 0.93, 0.85 to 1.02; adjusted HR 1.07, 0.87 to 1.31; Figure 2.)

**Figure 2.**
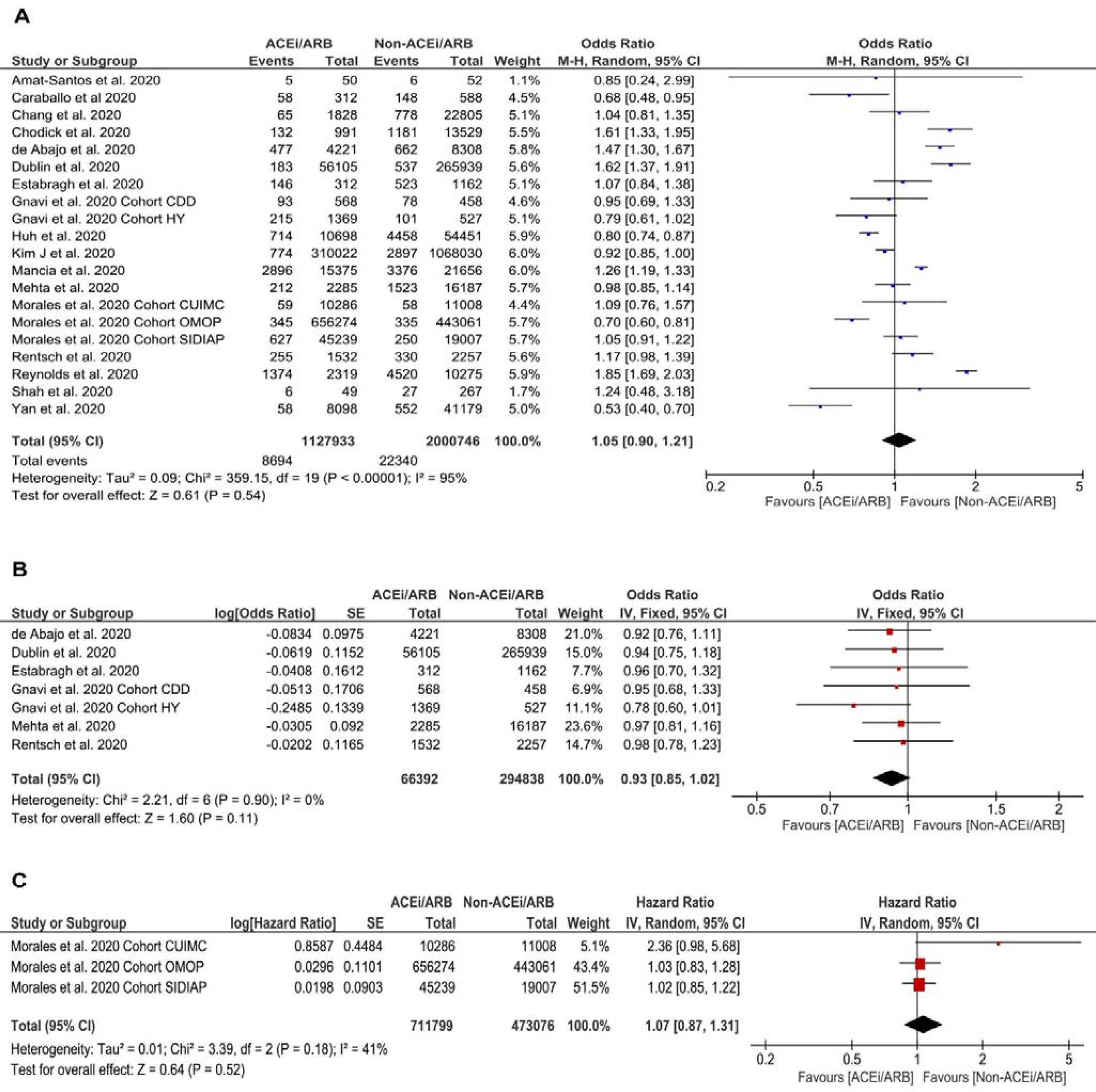
Pooled unadjusted OR (A) and adjusted OR (B) and adjusted HR (C) for the association of susceptibility of COVID-19 with RAS inhibitor use

### Use of RAS inhibitors and the severity of COVID-19

We assessed the Covid-19 case severity from 29 studies, including 18,959 COVID-19 patients, of which 3840 were severe cases (20.3%). Adjusted ORs were reported for 13 studies and adjusted HRs were reported for 1 study. The definitions of severe COVID-19 are listed in eTable 4 in the Supplement. There was no significant difference in severe case rate between RAS inhibitor users and non-users before (OR 1.05, 0.81 to 1.36) and after adjustment for confounders (OR 0.76, 0.52 to 1.12), especially age and gender (Figure 3).

**Figure 3.**
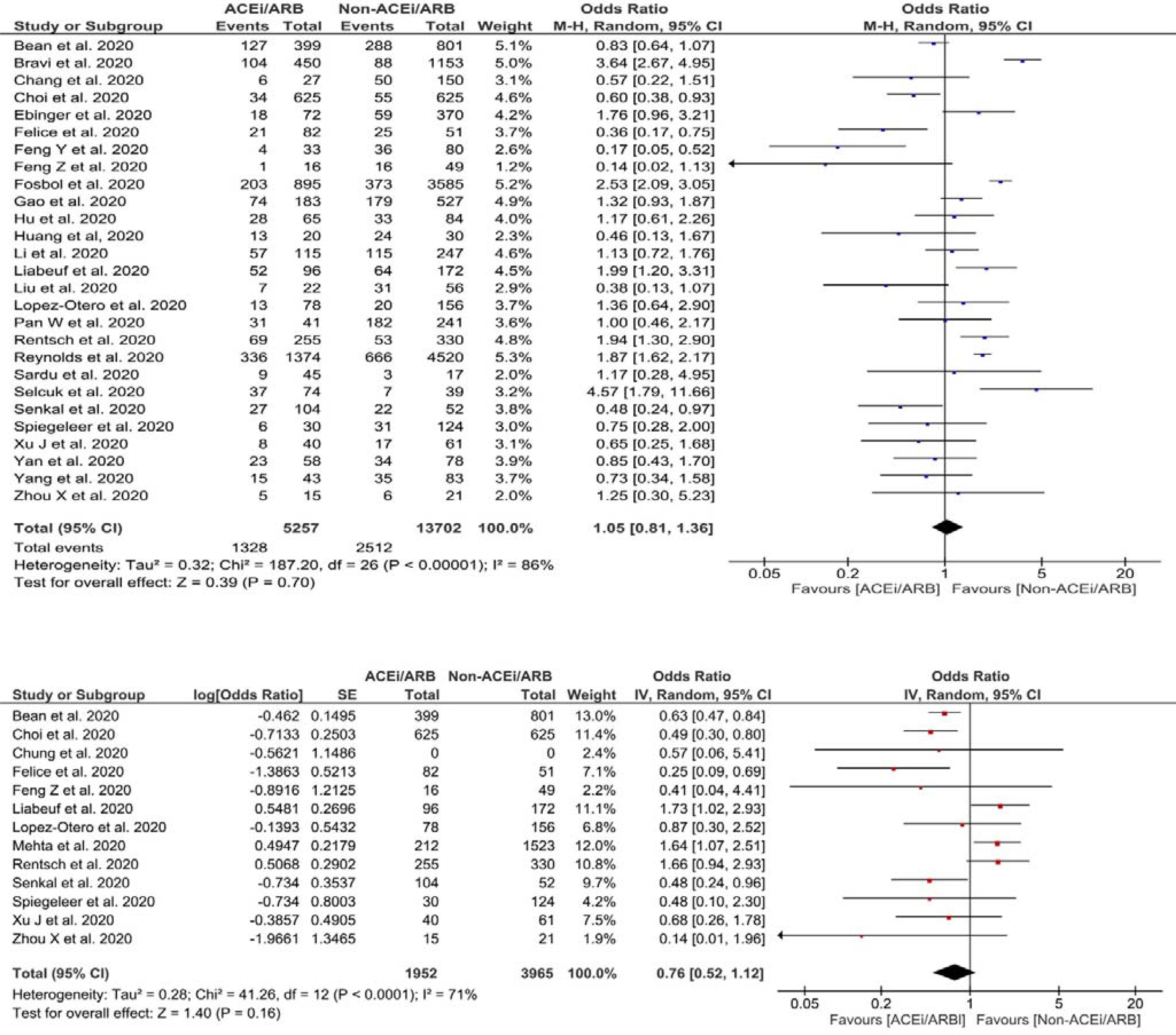
Pooled unadjusted (A) and adjusted (B) OR for the association of severe COVID-19 with RAS inhibitor use

### Use of RAS inhibitors and mortality from COVID-19

30 studies were included in the unadjusted analysis and 11 studies in the adjusted analysis of mortality from COVID-19 (8 with adjusted ORs and 3 with adjusted HRs). In 33,441 COVID-19 patients, 4234 deaths (12.7%) occurred. There was no significant difference in the mortality between RAS inhibitor users and non-users before (OR 1.12, 0.88 to 1.44) and after adjustment for age, gender and other covariates (OR 0.97, 0.77 to 1.23, Figure 4). Three studies reported adjusted HRs, for which the pooled HR showed no difference in mortality in RAS inhibitor users when compared with non-users in COVID-19 patients (HR 0.62, 0.34 to 1.14).

**Figure 4.**
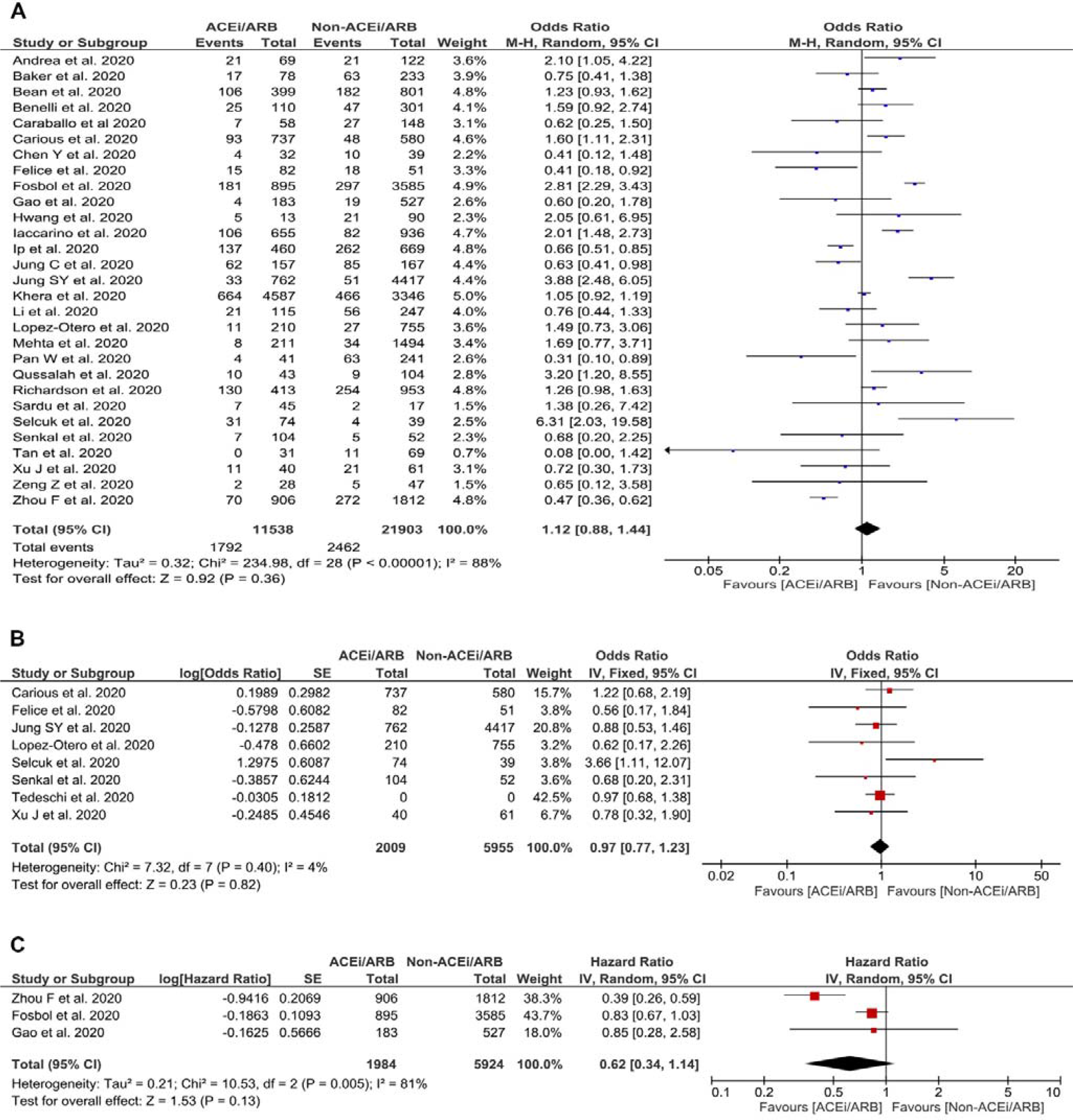
Pooled unadjusted OR (A) and adjusted OR (B) and adjusted HR (C) for the association of mortality of COVID-19 with RAS inhibitor use

## Subgroup analyses

### 1. A comparison of the association of ACEi versus ARB with susceptibility and severity of COVID-19

When ACEi and ARB were analyzed separately (eFigures 1 and 2 in the Supplement), before and after adjustment for confounders, there was no significant difference in the incidence of COVID-19 infection between ACEi users and non-users (unadjusted OR 0.94, 0.89 to 1.00 and adjusted OR 0.90, 0.79 to 1.04) or between ARB users and non-users (unadjusted OR 1.05, 1.00 to 1.11 and adjusted OR 1.12, 0.96 to 1.32). There was also no difference in the severe case rate between ACEi users and non-users (OR 0.93, 0.59 to 1.48), or between ARB users and non-users (OR 0.91, 0.71 to 1.17, eFigure 3 in the Supplement). When COVID-19 disease severity was subdivided into mild-to-moderate or severe cases, with 2 studies reporting data after adjustment, and using non-COVID-19 subjects as reference, (eFigure 4 in the Supplement), ACEi use was not associated with increased mild-to-moderate case severity (OR 0.97, 0.88 to 1.06), or severe case severity (OR 0.82, 0.67 to 1.01) and neither was ARB use associated with increased mild-to-moderate case severity (OR 0.96, 0.88 to 1.06) or severe case severity (OR 1.01, 0.67 to 1.50).

### 2. Impact of RAS inhibitor use in patients with hypertension on susceptibility to Covid-19 and disease severity

In the susceptibility analysis without adjustment, 4 studies reported data specifically in hypertensive patients (eFigure 5 in the Supplement). There was no difference in the incidence of COVID-19 infection between RAS inhibitor users and non-users (OR 1.13, 0.80 to 1.61) in patients with hypertension. Similar findings were observed after adjustment (OR 0.87, 0.73 to 1.04, eFigure 6 in the Supplement).

Seventeen studies reported unadjusted data on disease severity in hypertensive COVID-19 patients and there was no difference in the severe case rate between RAS inhibitor users and non-users (OR 0.82, 0.63 to 1.06). Similarly, in 6 studies that reported adjusted ORs, there was also no significant difference after adjustment, with a OR 0.64 (0.40 to 1.02) (eFigure 6 in the Supplement).

Ten studies reported mortality in hypertensive COVID-19 patients and there was no difference in mortality between RAS inhibitor users and non-users before (OR 0.83, 0.60 to 1.13, eFigure 5 in the Supplement) and after adjustment (OR 0.93, 0.61 to 1.42 and HR 0.61, 0.33 o 1.12, eFigure 6 in the Supplement)

### 3. Association of susceptibility of COVID-19 with use of RAS inhibitors in case-controlled studies

Eight case-controlled studies and eleven retrospective cohort studies reported the susceptibility data without adjustment, which showed no difference in the incidence of COVID-19 infection between RAS inhibitor users and non-users, with OR 0.95, 0.78 to 1.15 and OR 1.14, 0.90 to 1.43, respectively, eFigure 7 in the Supplement).

### 4. Association of severity of COVID-19 with use of RAS inhibitors in Chinese and non-Chinese cohort

There were 12 studies reporting the severity data in Chinese cohorts without adjustment (eFigure 8 in the Supplement), which showed no difference in the incidence of severe case rate between Chinese RAS inhibitor users and non-users before (OR 0.79, 0.57 to 1.09, eFigure 8 in the Supplement) and after adjustment (OR 0.54, 0.23 to 1.26, eFigure 9 in the Supplement). In 14 studies in the non-Chinese cohorts, RAS inhibitor users had a higher severity rate than non-users before adjustment for confounders (OR 1.41, 1.02 to 1.96, eFigure 8 in the Supplement), but after adjustment, the associations were not significant (OR 0.80, 0.53 to 1.20, eFigure 9 in the Supplement).

### 5. Association of susceptibility and severity of COVID-19 with hypertension status

After adjustment for confounders, hypertension was not associated with the increased susceptibility to COVID-19 (OR 1.09, 0.95 to 1.25). Hypertension was significantly associated with the increased Covid-19 severity rate (OR 1.44, 1.10 to 1.89) and increased mortality after adjustment (HR 2.14, 1.46 to 3.14; eFigure 10 in the Supplement). However, when we included recently published UK national data (n=17,278,392) which focused on the association between co-morbidities (including hypertension) and COVID-19 mortality but without RAS inhibitors data^8^, there was no significant association between hypertension and COVID-19 mortality, with a HR 1.53, 0.78 to 2.99 (eFigure 10 in the Supplement).

### Sensitivity Analyses and Risk of Bias

The main outcomes of this meta-analysis remained unaltered in the sensitivity analysis by excluding each individual study. Considering the retrospective and observational setting for all of these studies, the risk of bias was high (eTables 1-3 in the Supplement) and the I^2^ values were large in some pooled analyses. However, Funnel plots and Egger’s test indicated that there was no substantial publication bias (eFigures 11-13 in the Supplement).

## Discussion

In this systematic review and meta-analysis of observational and case-controlled studies, the use of RAS inhibitors was not associated with increased susceptibility to infection with SARS-CoV-2 and was not associated with increased risk of severe disease or mortality from COVID-19. Moreover, when ACEi or ARB were analyzed separately versus non-users of these drugs, there was no evidence that either treatment’s association with susceptibility, severe disease or death from COVID-19 was different from the other when compared to non-users. Similar results were obtained when specifically examining RAS inhibitor users versus non-use in people with hypertension, or when comparing Chinese and non-Chinese patient cohorts and when the analysis was restricted to case-controlled studies.

Due to the fact that the earliest studies characterizing co-morbidities most likely to be associated with poor outcomes from COVID-19, cited pre-existing hypertension as one of the most important,^9^ we also examined whether hypertension was independently associated with increased susceptibility to infection with SARS-CoV-2 and increased risk of severe disease or mortality in those infected. Mindful of how common hypertension is and its strong association with ageing, we had concerns that the Covid-19 risk associated with hypertension might have been exaggerated because the earliest reports had not been adjusted for potential confounding, especially age, the latter being a potent risk factor for death from COVID-19.^8^ Thus, it was important to examine the association between COVID-19 and hypertension independent of age, other demographics, and comorbidities. Our pooled studies with available data on hypertension (6 studies for susceptibility) showed no significant independent association between hypertension and susceptibility to SARS-CoV-2 infection. However, we did find a significant association between hypertension and case severity and mortality from Covid-19. Subsequently, we incorporated a new analysis by Williamson et al^8^, of the relationship between patient demographics and comorbidities (including hypertension) and Covid-19 mortality in 17 million people in the UK. The addition of this data to our analysis, attenuated the association between hypertension and mortality, however, the pooled results still revealed a trend towards an approximately 50% increased probability of an adverse outcome in patients with hypertension, although the corresponding confidence interval crossed the reference line. Interestingly, the report of Williamson et al^8^, reveals the complexity and the importance of confounding in the relationships between hypertension and Covid-19 outcomes. Notably that much of the risk association between hypertension and Covid-19 outcomes could be accounted for by age, diabetes and obesity, which are all independent risk factors for poorer outcomes and are commonly associated with hypertension. There was also an intriguing interaction between age and hypertension whereby hypertension was associated with increased risk of an adverse outcome from Covid-19 up to the age of 70 years but reduced risk beyond 70 years of age. This illustrates the complexity of interpreting associations for common comorbidities when there is often complex or unmeasured residual confounding from the many cardiac, vascular and metabolic comorbidities associated with hypertension., rather than attributing risk to an elevated blood pressure *per se*.

Although often considered similar in their RAS inhibiting effects, ACEi and ARBs block the RAS pathway at different levels which could theoretically differentially influence the level and activity of ACE2, the availability of angiotensin II at the AT-2 receptor and the angiotensin II cleavage product, angiotensin 1-7. Hypotheses have been presented to explain how these differential effects of ACEi and ARBs could influence the susceptibility to SARS-CoV-2 infection and/or severity and mortality from COVID-19. For the purposes of our analysis, many studies reported results with ACEi and ARB separately and we found that the pooled results of either ACEi or ARB showed no association with COVID-19 for any outcomes of interest. One study directly compared the effects of ACEi and ARB on mortality from COVID-19. By comparing 124 COVID-19 patients taking ACEi with 248 matched patients taking ARB, no significant difference in 28-day all-cause mortality were found between groups.^10^ Our findings suggest that there neither RAS-inhibition overall, or ACEi or ARB treatment individually are associated with increased risk from COVID-19.

The concept that ACEi or ARB might increase the risk of infection or severity of COVID-19 arose from the observation that ACE2 is the binding site that facilitates the cellular entry of SARS-CoV-2. This led to speculation that treatment with RAS inhibitor drugs might increase the cellular expression of ACE2 and thereby increase the risk of infection and severity of disease. This speculation cited previous experimental studies that had shown that RAS inhibitor could increase the expression of ACE2 in heart and kidney^2^, however, there was no data on the effect of RAS inhibitors on lung ACE2 expression in either animals or human. Moreover, the concept that RAS inhibitors increases ACE2 expression is now being contested. A recent study, Sama et al.^11^ showed that use of an ACEi or an ARB was not associated with higher plasma ACE2 concentrations. Whilst circulating ACE2 may not be a good indicator of tissue ACE2 levels, other recent evidence suggests that there is increasing expression of ACE2 with age in both human lungs and kidney but no association between the expression of ACE2 with either hypertension or RAS inhibiting drugs.^12^ Thus, the hypothesis that RAS inhibitors could increase risk from COVID-19 by increasing ACE2 expression is unsupported by the limited pathophysiological data available, or the more extensive clinical data from the cohort studies of patients included in this meta-analysis. This in turn, reinforces the recommendations made by medical societies, for patients to continue with their RAS inhibitor medications throughout this COVID-19 pandemic. ^3^

The present finding needs to be interpreted, mindful of the limitations of observational cohort studies. It is noteworthy that in most analyses, there is substantial risk of bias indicated by the high I^2^ value in both main and subgroup analyses. This was mainly attributable to the inherent weakness of observational studies and the inevitable possibility of confounding. The association of various outcomes or with prior pharmaceutical treatments are commonly influenced by numerous confounders, especially the indication for treatment, which often varies according to comorbidities. The fact that RAS inhibitor medications are recommended as a foundation treatment for hypertension or many of the comorbidities associated with increased risk of mortality from COVID-19, i.e. diabetes, cardiac disease and chronic kidney disease, and the fact that all of these conditions are more common with advancing age, highlights the obvious potential for confounding in the relationship between RAS inhibitor use and adverse outcomes from COVID-19. These confounding can only really be overcome by randomized allocations to treatments in controlled trials and such trials are ongoing.^13^

## Conclusions

In this systematic review and meta-analysis of all existing clinical data during the Covid-19 pandemic, we found that treatment with RAS inhibitors, specifically ACEi or ARBs, was not associated with increased risk of infection with SARS-CoV-2, or increased risk for the development of severe disease, or mortality from COVID-19, when compared to non-users of RAS inhibition. Our study suggests that ACEi or ARBs should not be discontinued solely on the basis of concerns about potential risks associated with COVID-19.

## Data Availability

not applicable

## Author Contribution

YZ and BW conceived the idea for the study and the analysis plan. YZ and SY performed the data extractions and analyses, supervised by YX. YZ and BW drafted the manuscript with critical input from SY and YZ.

