## supplementary for "Renin Angiotensin System Inhibition and Susceptibility and Outcomes from COVID-19: A Systematic Review and Meta-analysis of 69,200 COVID-19 Patients"

eAppendix 1. Search strategy

eTable 1. Quality of studies included for the outcome of susceptibility of COVID-19

eTable 2. Quality of studies included for the outcome of severity of COVID-19

eTable 3. Quality of studies included for the outcome of mortality of COVID-19

eTable 4. Definitions of severity of COVID-19 in each included study

eFigure 1. Pooled adjusted OR for uses of ACEi and ARB in associations with susceptibility of COVID-19 infection

eFigure 2. Pooled adjusted HR for uses of ACEi and ARB in associations with susceptibility of COVID-19 infection

eFigure 3. Pooled adjusted OR for ACEi and ARB in associations with severe COVID-19

eFigure 4. Pooled adjusted OR for ACEi and ARB in associations with susceptibility of COVID-19 in non-severe and severe patients

eFigure 5. Pooled unadjusted OR for RAS inhibitor use in associations with susceptibility (A) to and severity (B) and mortality (C) of COVID-19 in hypertensives

eFigure 6. Pooled adjusted OR or HR for RAS inhibitor use in associations with susceptibility (A) to and severity (B) and mortality (C/D) of COVID-19 in hypertensives

eFigure 7. Pooled unadjusted OR for the RAS inhibitor use in association with susceptibility of COVID-19 by study types

eFigure 8. Pooled unadjusted OR for RAS inhibitor use in association with severe COVID-19 in Chinese and non-Chinese

eFigure 9. Pooled adjusted OR for RAS inhibitor use in association with severe COVID-19 in Chinese and non-Chinese

eFigure 10. Pooled adjusted OR or HR for hypertension in association with susceptibility and severity and mortality of COVID-19

eFigure 11. Funnel plot assessing publication bias in the association of susceptibility of COVID-19 with use of RAS inhibitors

eFigure 12. Funnel plot assessing publication bias in the association of severe COVID-19 with use of RAS inhibitors

eFigure 13. Funnel plot assessing publication bias in the association of mortality of COVID-19 with use of RAS inhibitors

**eAppendix 1. Search strategy**

1.1. PubMed:

#1 (((((coronavirus disease 2019) OR 2019 novel coronavirus disease) OR COVID-19) OR Severe acute respiratory syndrome coronavirus 2) OR SARS-CoV-2) OR 2019-nCoV

#2 (Renin-Angiotensin System) OR Renin-Angiotensin-Aldosterone System)

#3 (Angiotensin Converting Enzyme Inhibitors) OR ACE Inhibitors) OR Angiotensin-Converting Enzyme Antagonists

#4 (Angiotensin II type 1 Receptor Blockers) OR (Angiotensin II Type 1 Receptor Antagonists)

#5((“2020/01/01”[Date-Publication]:“2020/07/20”[Date-Publication])) AND English[Language]

#6 #1 AND ((#2 OR #3) OR #4) AND #5

1.2. Web of science

#1 TS=(coronavirus disease 2019 OR 2019 novel coronavirus disease OR COVID-19 OR Severe acute respiratory syndrome coronavirus 2 OR SARS-CoV-2 OR 2019-nCoV)

Datasets=WOS, CSCD, DIIDW, KJD, MEDLINE, RSCI, SCIELO

Timespan=Year to date

Search language=English

#2 TS=(Renin-Angiotensin System OR Renin-Angiotensin-Aldosterone System)

Datasets=WOS, CSCD, DIIDW, KJD, MEDLINE, RSCI, SCIELO

Timespan=Year to date

Search language=English

#3 TS=(Angiotensin Converting Enzyme Inhibitors OR ACE Inhibitors OR Angiotensin-Converting Enzyme Antagonists)

Datasets=WOS, CSCD, DIIDW, KJD, MEDLINE, RSCI, SCIELO

Timespan=Year to date

Search language=English

#4 TS=(Angiotensin II type 1 Receptor Blockers OR Angiotensin II Type 1 Receptor Antagonists)

Datasets=WOS, CSCD, DIIDW, KJD, MEDLINE, RSCI, SCIELO

Timespan=Year to date

Search language=English

#5 #2 OR #3

#6 #4 OR #5

#7 #1 AND #6

1.3. Scopus

TITLE-ABS-KEY(“coronavirus disease 2019” OR “2019 novel coronavirus disease” OR “COVID-19” OR “Severe acute respiratory syndrome coronavirus 2” OR “SARS-CoV-2” OR 2019-nCoV) AND TITLE-ABS-KEY(“Renin-Angiotensin System” OR “Renin-Angiotensin-Aldosterone System” OR “Angiotensin Converting Enzyme Inhibitors” OR “ACE Inhibitors” OR “Angiotensin-Converting Enzyme Antagonists” OR “Angiotensin II type 1 Receptor Blockers” OR “Angiotensin II type 1 Receptor Antagonists”) AND LANGUAGE(English) AND PUBYEAR AFT 2019

1.4 OVID Embase

#1 (coronavirus disease 2019 or 2019 novel coronavirus disease or COVID-19 or Severe acute respiratory syndrome coronavirus 2 or SARS-CoV-2 or 2019-nCoV).af.

#2 (Renin-Angiotensin System or Renin-Angiotensin-Aldosterone System).af.

#3 (Angiotensin Converting Enzyme Inhibitors or ACE Inhibitors or Angiotensin-Converting Enzyme Antagonists).af.

#4 (Angiotensin II type 1 Receptor Blockers or Angiotensin II Type 1 Receptor Antagonists).af.

#5 2 or 3

#6 4 or 5

#7 1 and 6

#8 limit to (english language an d yr="2020 -Current")

**eTable 1. Quality of studies included for the outcome of susceptibility of COVID-19**

| **Case control studies** | | | | | | | | | | |
| --- | --- | --- | --- | --- | --- | --- | --- | --- | --- | --- |
| Author year | Selection | | | | Comparability | | Exposure | | | Total quality score |
|  | Is the case definition adequate? | Representativeness of the cases | Selection of Controls | Definition of Controls | Most important factor | Second important factor | Ascertainment of exposure | Same method of ascertainment for cases and controls | Non-Response rate |  |
| Chang et al. 2020 | ☆ | ☆ | ☆ | ☆ |  |  | ☆ | ☆ |  | 6 |
| de Abajo et al. 2020 | ☆ | ☆ | ☆ |  | ☆ | ☆ | ☆ | ☆ | ☆ | 8 |
| Fosbol et al. 2020 | ☆ |  | ☆ | ☆ | ☆ | ☆ | ☆ | ☆ |  | 7 |
| Gnavi et al. 2020 | ☆ |  | ☆ |  | ☆ |  |  | ☆ | ☆ | 5 |
| Gnavi et al. 2020 | ☆ |  | ☆ |  | ☆ |  |  | ☆ | ☆ | 5 |
| Huh et al. 2020 | ☆ | ☆ | ☆ | ☆ |  |  | ☆ | ☆ | ☆ | 7 |
| Mancia et al. 2020 | ☆ | ☆ | ☆ |  |  |  | ☆ | ☆ | ☆ | 6 |
| Yan H et al.2020 | ☆ | ☆ | ☆ |  |  |  | ☆ | ☆ | ☆ | 6 |
| **Cohort studies** | | | | | | | | | | |
| Author year | Selection | | | | Comparability | | Exposure | | | Total quality score |
|  | Representativeness of the exposed cohort | Selection of the non-exposed cohort | Ascertainment of exposure | Demonstration that outcome of interest was not present at start of study | Most important factor | Second important factor | Assessment of outcome | Was follow-up long enough for outcomes to occur | Adequacy of follow up of cohorts |  |
| Amat-Santos et al. 2020 | ☆ | ☆ | ☆ | ☆ |  |  |  | ☆ | ☆ |  |
| Caraballo et al. 2020 | ☆ | ☆ | ☆ | ☆ |  |  | ☆ |  |  | 5 |
| Chodick et al 2020 | ☆ | ☆ | ☆ | ☆ |  |  | ☆ |  |  | 5 |
| Dublin et al. 2020 | ☆ | ☆ | ☆ | ☆ | ☆ | ☆ |  | ☆ | ☆ | 8 |
| Estabragh et al. 2020 | ☆ | ☆ |  | ☆ | ☆ | ☆ | ☆ |  |  | 7 |
| Kim J et al. 2020 | ☆ | ☆ | ☆ | ☆ |  |  |  |  |  | 4 |
| Mehta et al. 2020 | ☆ | ☆ | ☆ | ☆ | ☆ | ☆ | ☆ |  |  | 7 |
| Morales et al. 2020 (SIDIAP) | ☆ | ☆ | ☆ | ☆ | ☆ | ☆ |  |  |  | 6 |
| Morales et al. 2020 (VA-OMOP) | ☆ | ☆ | ☆ | ☆ | ☆ | ☆ |  |  |  | 6 |
| Morales et al. 2020 (CUIMC) | ☆ | ☆ | ☆ | ☆ | ☆ | ☆ |  |  |  | 6 |
| Rentsch et al. 2020 | ☆ | ☆ | ☆ | ☆ | ☆ | ☆ | ☆ |  |  | 7 |
| Reynolds et al. 2020 | ☆ | ☆ | ☆ | ☆ |  |  | ☆ |  |  | 5 |
| Shah et al. 2020 | ☆ | ☆ | ☆ | ☆ |  |  | ☆ |  |  | 5 |

**eTable 2. Quality of studies included for the outcome of severity of COVID-19**

| **Case control study** | | | | | | | | | | |
| --- | --- | --- | --- | --- | --- | --- | --- | --- | --- | --- |
| Author year | Selection | | | | Comparability | | Exposure | | |  |
|  | Is the case definition adequate? | Representativeness of the cases | Selection of Controls | Definition of Controls | Most important factor | Second important factor | Ascertainment of exposure | Same method of ascertainment for cases and controls | Non-Response rate | Total quality score |
| Bravi et al. 2020 | **☆** | **☆** | **☆** |  |  |  | **☆** | **☆** | **☆** | 6 |
| Chang et al. 2020 | **☆** |  | **☆** |  |  |  | **☆** | **☆** |  | 4 |
| **Cohort study** |  |  |  |  |  |  |  |  |  |  |
| Author year | Selection | | | | Comparability | | Exposure | | |  |
|  | Representativeness of the exposed cohort | Selection of the non-exposed cohort | Ascertainment of exposure | Demonstration that outcome of interest was not present at start of study | Most important factor | Second important factor | Assessment of outcome | Was follow-up long enough for outcomes to occur | Adequacy of follow up of cohorts | Total quality score |
| Bean et al. 2020 | **☆** | **☆** | **☆** | **☆** | **☆** | **☆** | **☆** |  | **☆** | 8 |
| Choi et al. 2020 | **☆** | **☆** | **☆** | **☆** | **☆** | **☆** | **☆** |  |  | 7 |
| Chung et al. 2020 |  | **☆** | **☆** | **☆** | **☆** | **☆** |  |  |  | 5 |
| Ebinger et al. 2020 | **☆** | **☆** | **☆** |  |  |  | **☆** |  |  | 4 |
| Felice et al. 2020 |  | **☆** | **☆** |  | **☆** | **☆** | **☆** |  |  | 5 |
| Feng Y et al. 2020 | **☆** | **☆** | **☆** |  |  |  | **☆** |  |  |  |
| Feng Z et al. 2020 |  | **☆** | **☆** |  | **☆** | **☆** | **☆** |  |  | 5 |
| Fosbol et al. 2020 | **☆** |  | **☆** | **☆** | **☆** | **☆** | **☆** | **☆** |  | 7 |
| Gao et al. 2020 |  | **☆** | **☆** |  |  |  |  | **☆** | **☆** | 5 |
| Hu et al. 2020 |  | **☆** | **☆** | **☆** |  |  |  |  |  | 3 |
| Huang et al. 2020 |  | **☆** | **☆** |  |  |  | **☆** |  |  |  |
| Li et al. 2020 |  | **☆** | **☆** |  |  |  | **☆** |  |  | 3 |
| Liabeuf et al. 2020 |  | **☆** | **☆** | **☆** | **☆** | **☆** | **☆** |  |  | 6 |
| Liu et al. 2020 |  | **☆** | **☆** |  |  |  | **☆** |  |  | 3 |
| Lopez-Otero et al. 2020 |  | **☆** | **☆** | **☆** | **☆** | **☆** | **☆** |  |  | 6 |
| Mehta et al. 2020 | **☆** | **☆** | **☆** | **☆** | **☆** | **☆** | **☆** |  |  | 7 |
| Pan W et al. 2020 |  | **☆** | **☆** | **☆** |  |  | **☆** |  |  | 4 |
| Rentsch et al. 2020 |  | **☆** | **☆** | **☆** | **☆** | **☆** | **☆** |  |  | 6 |
| Reynolds et al. 2020 | **☆** | **☆** | **☆** | **☆** | **☆** | **☆** | **☆** |  |  | 7 |
| Sardu et al. 2020 |  | **☆** | **☆** |  |  |  | **☆** |  | **☆** | 4 |
| Selcuk et al. 2020 |  | **☆** | **☆** | **☆** |  |  | **☆** | **☆** | **☆** | 6 |
| Senkal et al. 2020 |  | **☆** | **☆** | **☆** | **☆** | **☆** | **☆** | **☆** | **☆** | 8 |
| Spiegeleer et al. 2020 |  | **☆** |  | **☆** | **☆** | **☆** | **☆** |  |  | 5 |
| Xu J et al. 2020 |  | **☆** | **☆** | **☆** | **☆** |  |  | **☆** |  | 5 |
| Yan H et al. 2020 | **☆** | **☆** | **☆** | **☆** | **☆** |  | **☆** |  |  | 6 |
| Yang et al. 2020 |  | **☆** | **☆** |  | **☆** | **☆** | **☆** |  |  | 5 |
| Zhou X et al. 2020 |  | **☆** | **☆** |  |  |  | **☆** |  |  | 3 |

**eTable 3. Quality of studies included for the outcome of mortality of COVID-19**

| Author year | Selection | | | | Comparability | | Exposure | | | Total quality score |
| --- | --- | --- | --- | --- | --- | --- | --- | --- | --- | --- |
|  | Representativeness of the exposed cohort | Selection of the non-exposed cohort | Ascertainment of exposure | Demonstration that outcome of interest was not present at start of study | Most important factor | Second important factor | Assessment of outcome | Was follow-up long enough for outcomes to occur | Adequacy of follow up of cohorts |  |
| Andrea et al. 2020 |  | **☆** | **☆** | **☆** | **☆** | **☆** | **☆** | **☆** | **☆** | 8 |
| Baker et al. 2020 |  |  | **☆** |  |  |  |  | **☆** |  | 2 |
| Bean et al. 2020 | **☆** | **☆** | **☆** | **☆** |  |  | **☆** |  | **☆** | 6 |
| Benelli et al. 2020 |  |  | **☆** |  |  |  | **☆** |  |  | 2 |
| Caraballo et al. 2020 |  | **☆** | **☆** | **☆** |  |  | **☆** |  |  | 4 |
| Carious et al. 2020 |  | **☆** | **☆** | **☆** | **☆** | **☆** | **☆** |  |  | 6 |
| Chen Y et al. 2020 |  |  |  | **☆** |  |  | **☆** |  |  | 2 |
| Felice et al. 2020 |  | **☆** | **☆** | **☆** | **☆** | **☆** | **☆** |  |  | 6 |
| Fosbol et al. 2020 | **☆** | **☆** | **☆** | **☆** | **☆** | **☆** | **☆** | **☆** | **☆** | 9 |
| Gao et al. 2020 |  | **☆** | **☆** | **☆** | **☆** | **☆** | **☆** | **☆** | **☆** | 8 |
| Hwang et al. 2020 |  |  | **☆** | **☆** |  |  | **☆** |  | **☆** | 4 |
| Iaccarino et al. 2020 |  | **☆** | **☆** |  |  |  | **☆** |  |  | 3 |
| Ip et al. 2020 |  |  |  | **☆** |  |  | **☆** |  |  | 2 |
| Jung C et al. 2020 |  |  |  | **☆** | **☆** | **☆** | **☆** | **☆** |  | 5 |
| Jung SY et al. 2020 | **☆** | **☆** | **☆** | **☆** | **☆** | **☆** | **☆** |  |  | 7 |
| Khera et al. 2020 |  | **☆** | **☆** | **☆** |  |  | **☆** | **☆** | **☆** | 6 |
| Li et al. 2020 |  | **☆** | **☆** | **☆** |  |  | **☆** |  |  | 4 |
| Lopez-Otero et al. 2020 |  | **☆** | **☆** | **☆** | **☆** | **☆** | **☆** |  |  | 6 |
| Mehta et al. 2020 | **☆** | **☆** | **☆** | **☆** |  |  | **☆** | **☆** | **☆** | 7 |
| Pan W et al. 2020 |  | **☆** | **☆** | **☆** |  |  | **☆** |  |  | 4 |
| Qussalah et al 2020 |  | **☆** | **☆** | **☆** |  |  | **☆** | **☆** | **☆** | 6 |
| Richardson et al. 2020 |  | **☆** | **☆** | **☆** |  |  | **☆** | **☆** | **☆** | 6 |
| Sardu et al. 2020 |  | **☆** | **☆** | **☆** |  |  | **☆** |  | **☆** | 5 |
| Selcuk et al. 2020 |  | **☆** | **☆** | **☆** |  |  | **☆** | **☆** | **☆** | 6 |
| Senkal et al. 2020 |  | **☆** | **☆** | **☆** | **☆** | **☆** | **☆** | **☆** | **☆** | 8 |
| Tan et al. 2020 |  | **☆** |  | **☆** |  |  | **☆** |  |  | 3 |
| Tedeschi et al. 2020 |  | **☆** |  | **☆** | **☆** | **☆** | **☆** |  |  | 5 |
| Xu J et al. 2020 |  | **☆** | **☆** | **☆** | **☆** |  | **☆** |  |  | 5 |
| Zeng Z et al. 2020 |  | **☆** |  | **☆** |  |  | **☆** |  |  | 3 |
| Zhou F et al. 2020 |  | **☆** | **☆** | **☆** | **☆** | **☆** | **☆** | **☆** | **☆** | 8 |

**eTable 4. Definitions of severity of COVID-19 in each included study**

| **Studies** | **Definitions of Severity** |
| --- | --- |
| Bean et al. 2020 | Death or admission to a critical care unit within 21 day of symptom onset |
| Bravi et al. 2020 | Admission in an intensive care unit and/or causing death |
| Chang et al. 2020 | Admitted to an intensive care unit or were intubated within 14 days of their first positive SARS-CoV-2 PCR test |
| Choi et al. 2020 | patients with one of the followings were considered as a severe infection case: 1) respiratory failure requiring mechanical ventilation, 2) organ failure requiring admission to the intensive care unit (ICU), 3) organ failure requiring continuous renal-replacement therapy (CRRT), or 4) organ failure  requiring extracorporeal membrane oxygenation (ECMO) treatment. |
| Chung et al. 2020 | composite outcome of acute respiratory distress syndrome, septic shock, intensive care unit care, and mortality within 28 days |
| Ebinger et al. 2020 | Admitted to ICU |
| Felice et al. 2020 | Admission to ICU/sICU |
| Feng Y et al. 2020 | the guidelines by the National Health Commission of China |
| Feng Z et al. 2020 | the guidelines by the National Health Commission of China |
| Fosbol et al. 2020 | ICU admission |
| Gao et al. 2020 | the guidelines by the National Health Commission of China |
| Hu et al. 2020 | the guidelines by the National Health Commission of China |
| Huang et al. 2020 | the guidelines by the National Health Commission of China |
| Li et al. 2020 | the guidelines by the National Health Commission of China |
| Liabeuf et al. 2020 | ICU admission or death before ICU admission. |
| Liu et al. 2020 | the guidelines by the National Health Commission of China |
| Lopez-Otero et al. 2020 | ICU admission |
| Mehta et al. 2020 | ICU admission |
| Pan W et al. 2020 | the guidelines by the National Health Commission of China |
| Rentsch et al. 2020 | admission to ICU |
| Reynolds et al. 2020 | admission to the intensive care unit (ICU), the use of invasive or noninvasive mechanical ventilation, or death. |
| Sardu et al. 2020 | ICU admission |
| Selcuk et al. 2020 | ICU admission |
| Spiegeleer et al. 2020 | long-stay hospital 131 admission (length of stay≥7 days) or death (at nursing home or hospital) within 14 days of disease onset |
| Spiegeleer et al. 2020 | serious COVID-19, i.e. long-stay hospital admission (length of stay≥7 days) or death (at nursing home or hospital) |
| Xu J et al. 2020 | ICU admission |
| Yan H et al. 2020 | the guidelines by the National Health Commission of China |
| Yang et al. 2020 | the guidelines by the National Health Commission of China |
| Zhou X et al. 2020 | Poor prognosis: Critical care and transfer to high level hospital |

Notes: The details of the guidelines by the National Health Commission of China, as below:

1. Mild type

The clinical symptoms are mild with no abnormal radiological findings.

2. Moderate type

Fever, cough and other symptoms are presented with pneumonia on chest computed

tomography.

3. Severe type

The disease is classified as severe if one of the following conditions is met:

(1) Respiratory distress, respiratory rate ≥ 30 per min;

(2) Oxygen saturation on room air at rest ≤ 93%;

(3) Partial pressure of oxygen in arterial blood / fraction of inspired oxygen ≤ 300 mmHg.

4. Critical type

One of the following conditions has to be met:

(1) Respiratory failure occurs and mechanical ventilation is required;

(2) Shock occurs;

(3) Patients with other organ dysfunction needing intensive care unit monitoring treatment.

In the included studies in our meta-analysis, mild and moderate types were thought as non-severe group, however, severe and critical types were treated as severe group.

**eFigure 1. Pooled adjusted OR for uses of ACEi and ARB in associations with susceptibility of COVID-19 infection**

**ACEi**

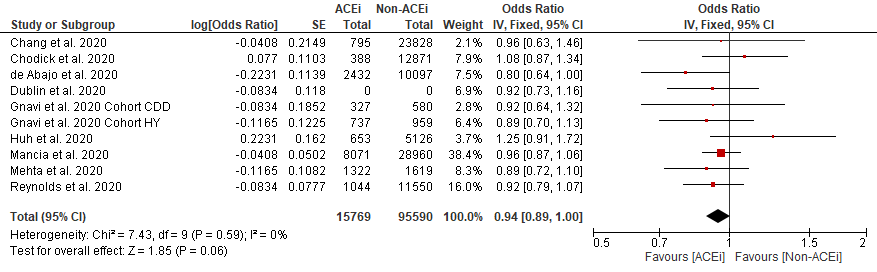

**ARB**

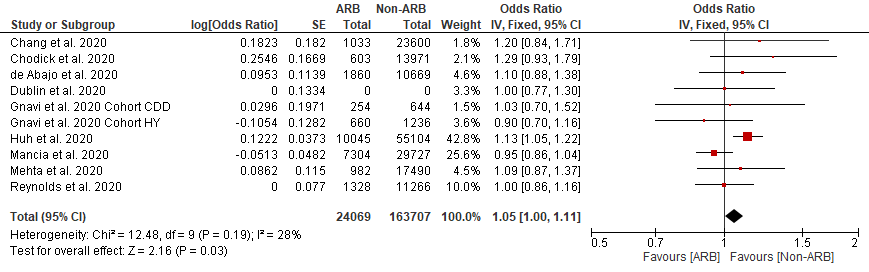

**eFigure 2. Pooled adjusted HR for uses of ACEi and ARB in associations with susceptibility of COVID-19 infection**

**ACEi**

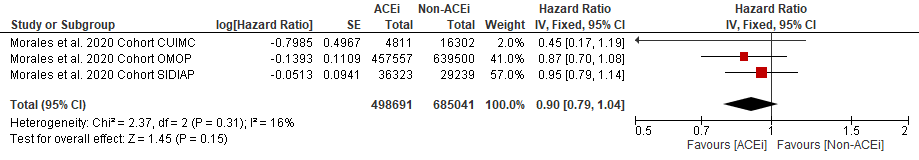

**ARB**

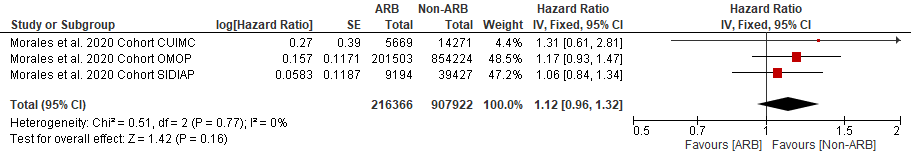

**eFigure 3. Pooled adjusted OR for ACEi and ARB in associations with severe COVID-19**

**ACEi**

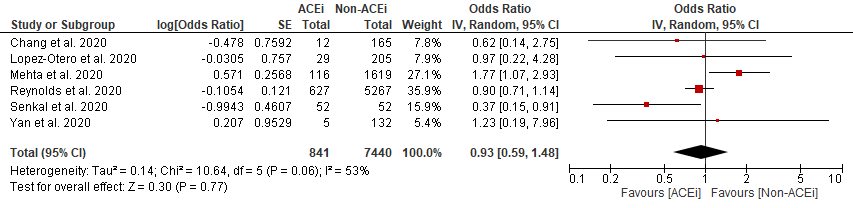

**ARB**

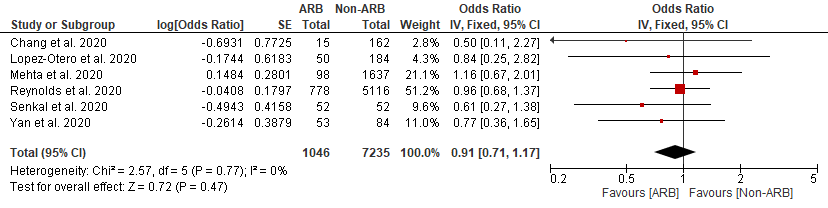

**eFigure 4. Pooled adjusted OR for ACEi and ARB in associations with susceptibility of COVID-19 in non-severe and severe patients**

**ACEi**

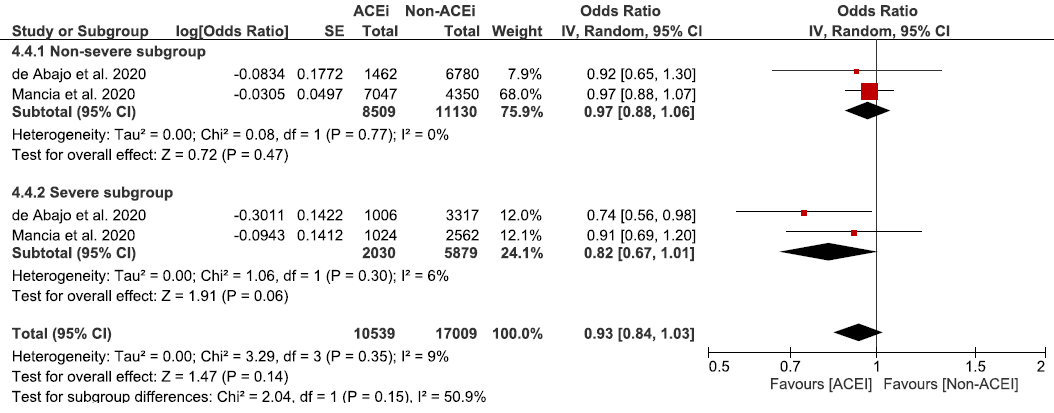

**ARB**

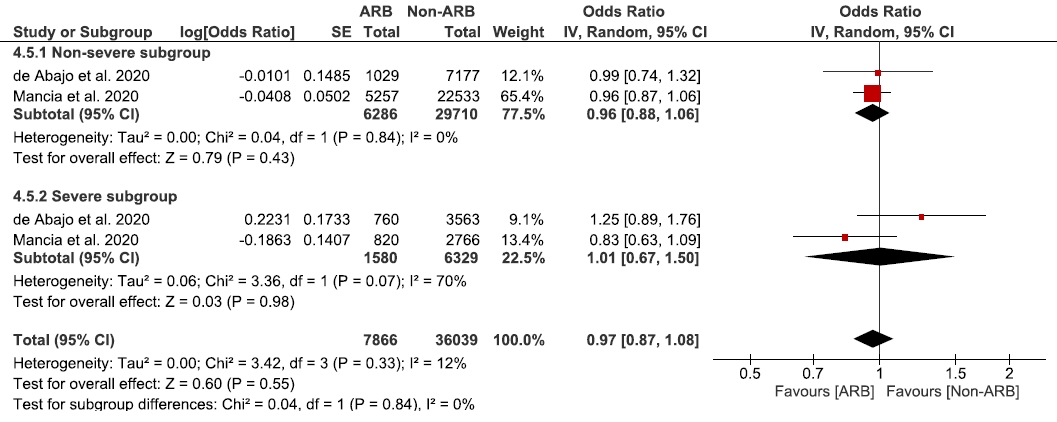

**eFigure 5. Pooled unadjusted OR for RAS inhibitor use in associations with susceptibility (A) to and severity (B) and mortality (C) of COVID-19 in hypertensives**

**A**

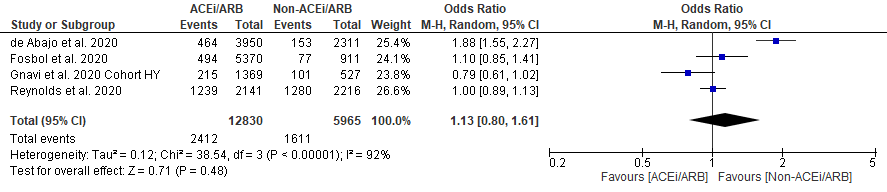

**B**

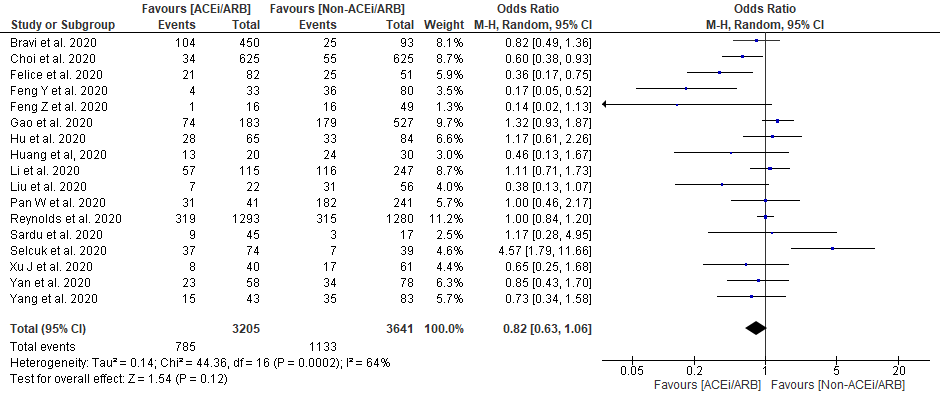

**C**

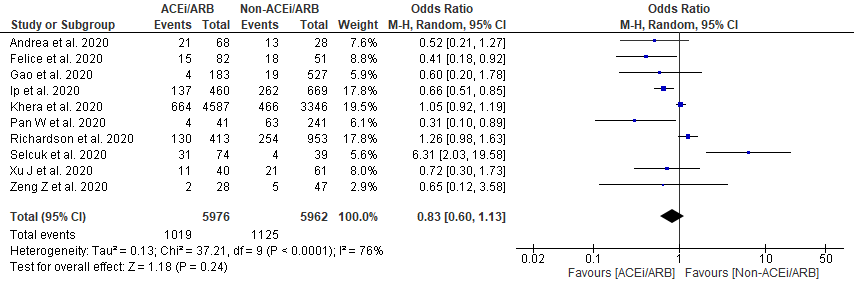

**eFigure 6. Pooled adjusted OR or HR for RAS inhibitor use in associations with susceptibility (A) to and severity (B) and mortality (C/D) of COVID-19 in hypertensives**

**A**

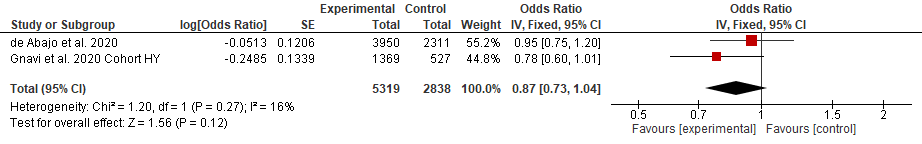

B

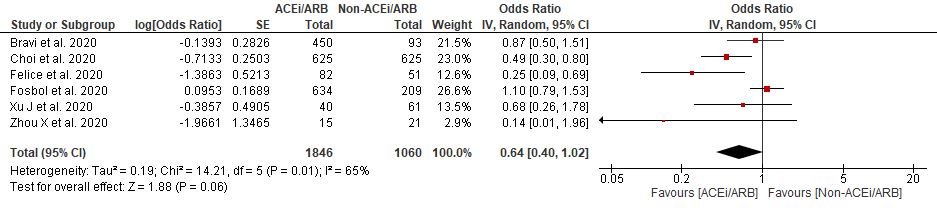

**C**

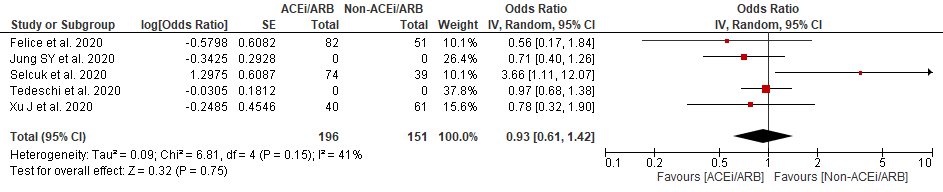

**D**

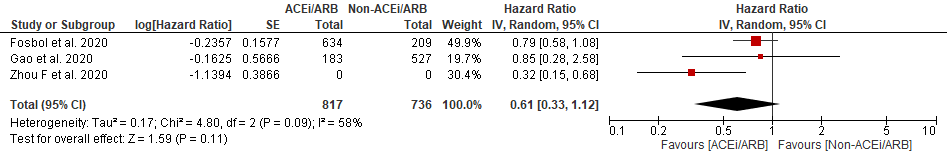

**eFigure 7. Pooled unadjusted OR for the RAS inhibitor use in association with susceptibility of COVID-19 by study types**

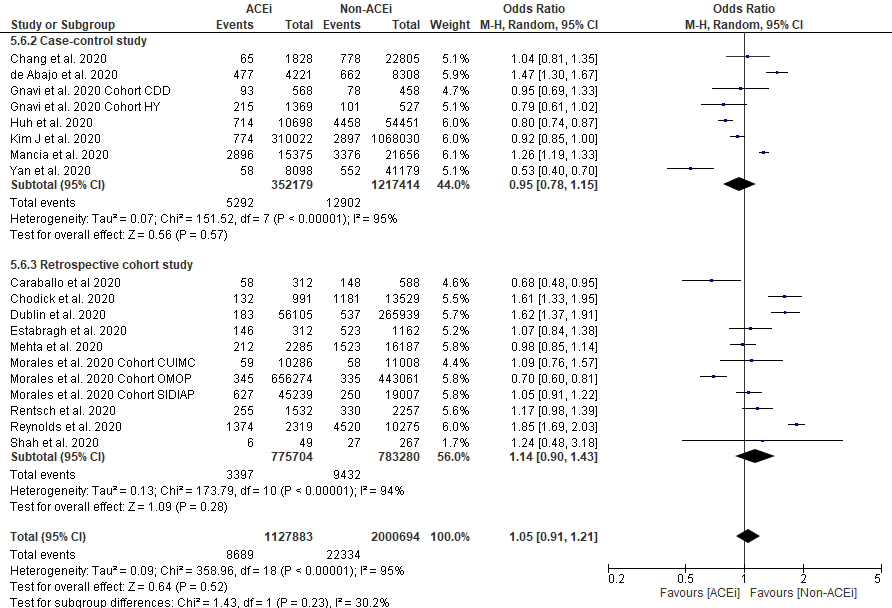

**eFigure 8. Pooled unadjusted OR for RAS inhibitor use in association with severe COVID-19 in Chinese and non-Chinese**

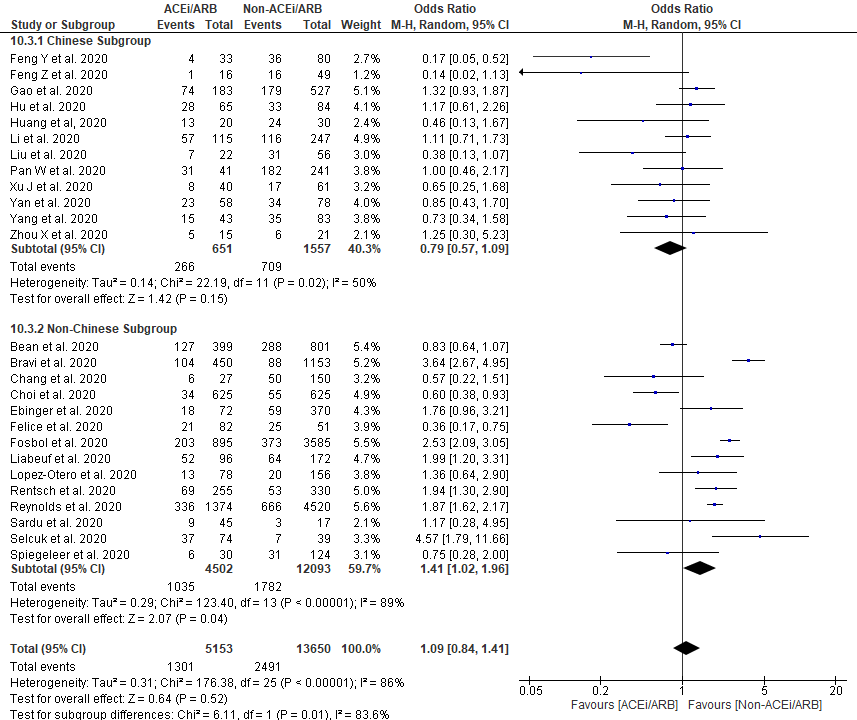

**eFigure 9. Pooled adjusted OR for RAS inhibitor use in association with severe COVID-19 in Chinese and non-Chinese**

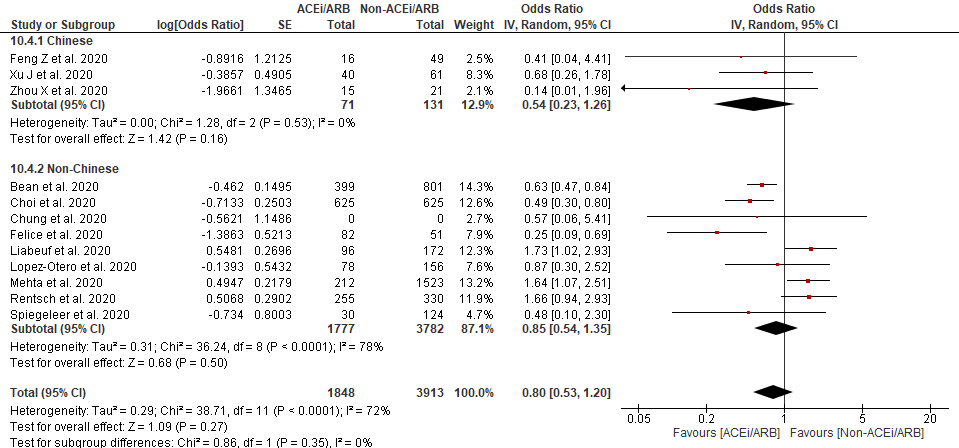

**eFigure 10. Pooled adjusted OR or HR for hypertension in association with susceptibility and severity and mortality of COVID-19**

**A**

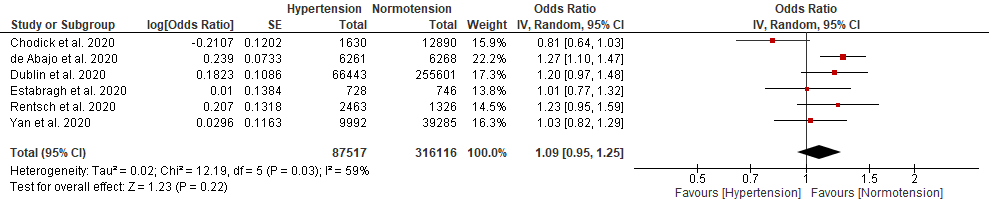

**B**

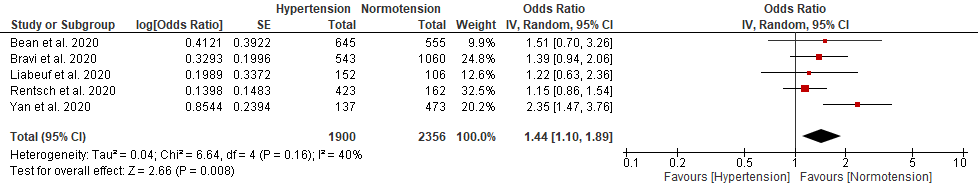

**C**

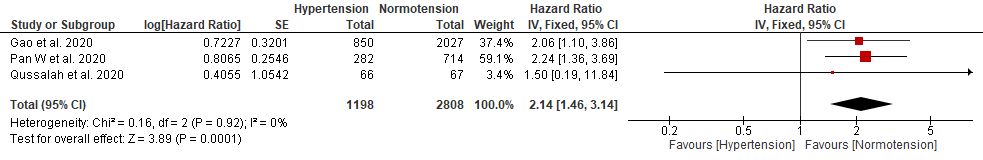

**D**

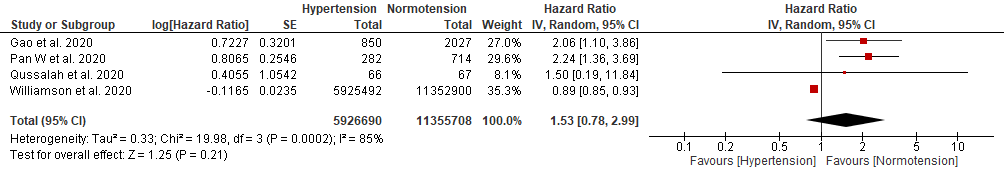

A, pooled adjusted OR for susceptibility; B, pooled adjusted OR for severity; C, pooled adjusted HR for mortality; D, pooled adjusted HR for mortality following the addition of a very large, recently published UK national cohort study [Williamson EJ, et al. Nature. 2020;10.1038/s41586-020-2521-4. doi:10.1038/s41586-020-2521-4].

**eFigure 11. Funnel plot assessing publication bias in the association of susceptibility of COVID-19 with use of RAS inhibitors**

Unadjusted model (P_Egger‘s test_=0.44)

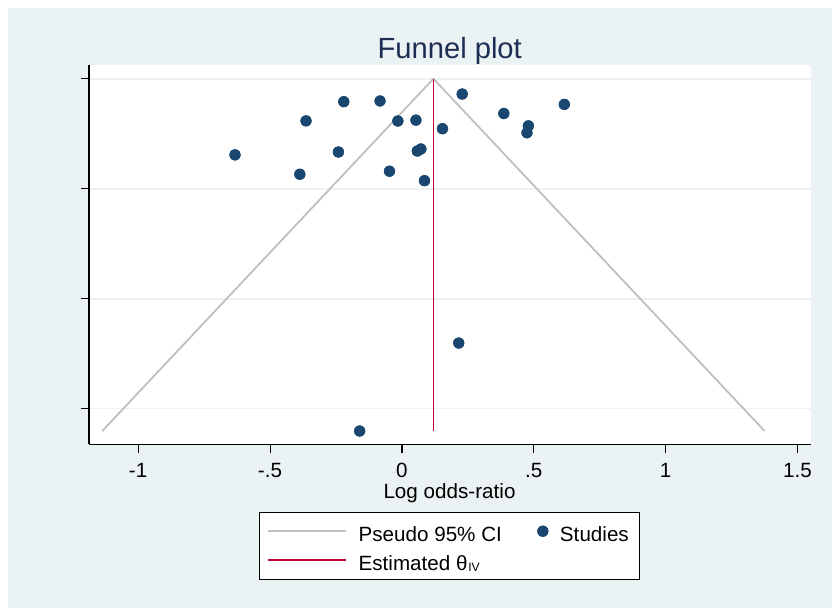

Adjusted OR model (P_Egger‘s test_=0.91)

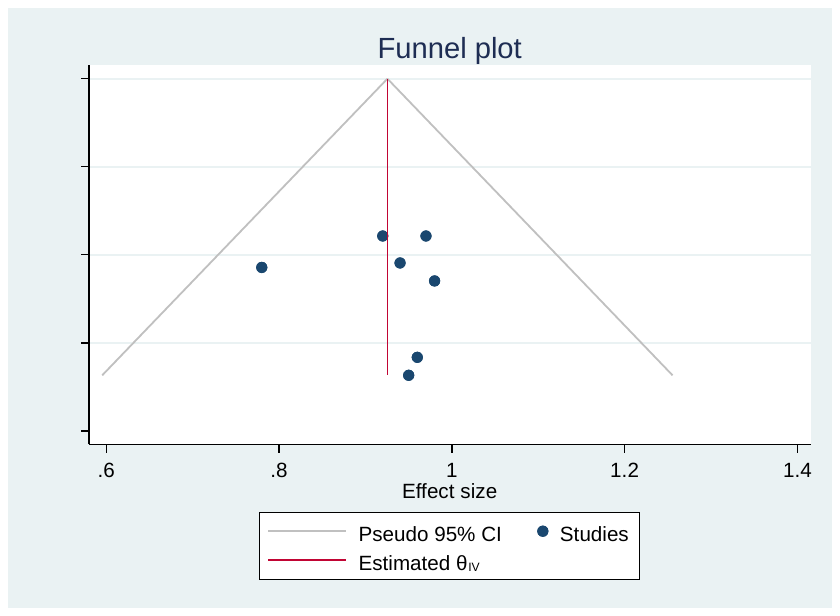

Adjusted HR model (P_Egger‘s test_=0.47)

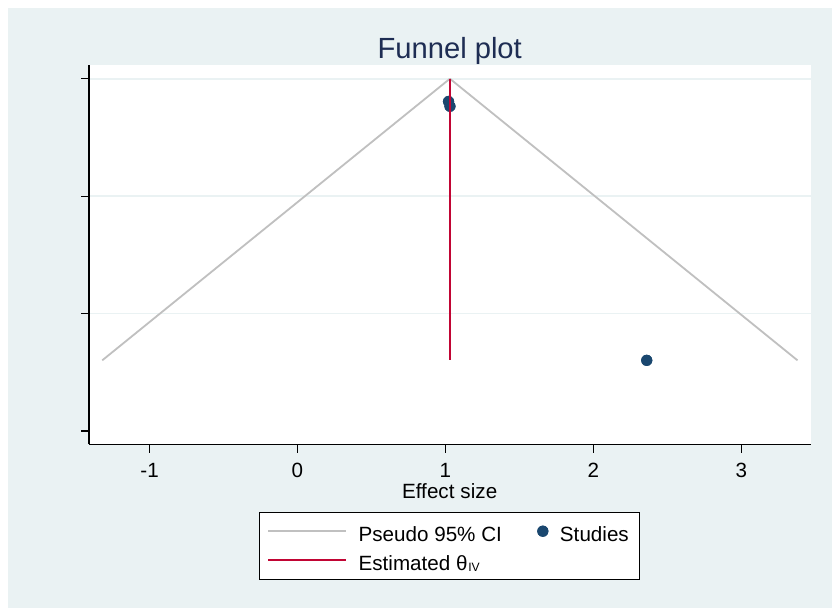

**eFigure 12. Funnel plot assessing publication bias in the association of severe COVID-19 with use of RAS inhibitors**

unadjusted model (P_Egger‘s test_<0.01)

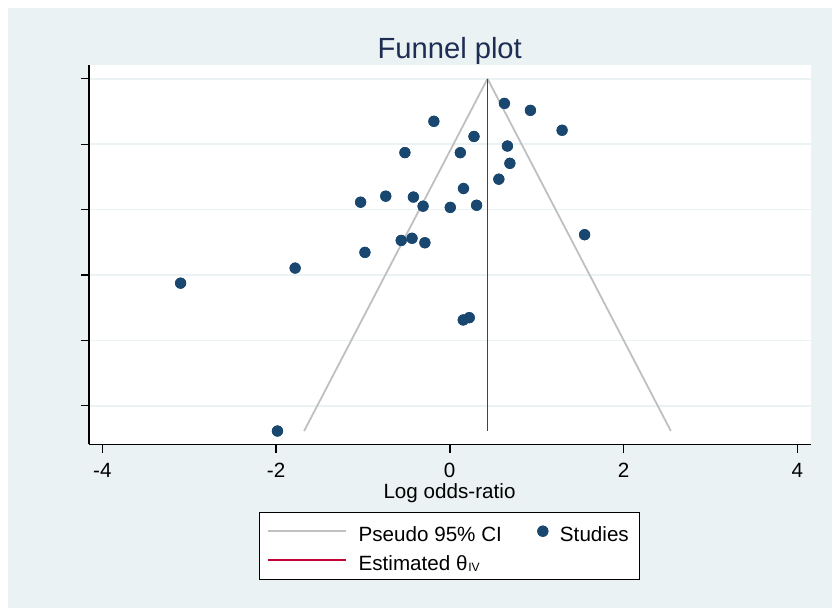

Adjusted model (P_Egger‘s test_=0.17)

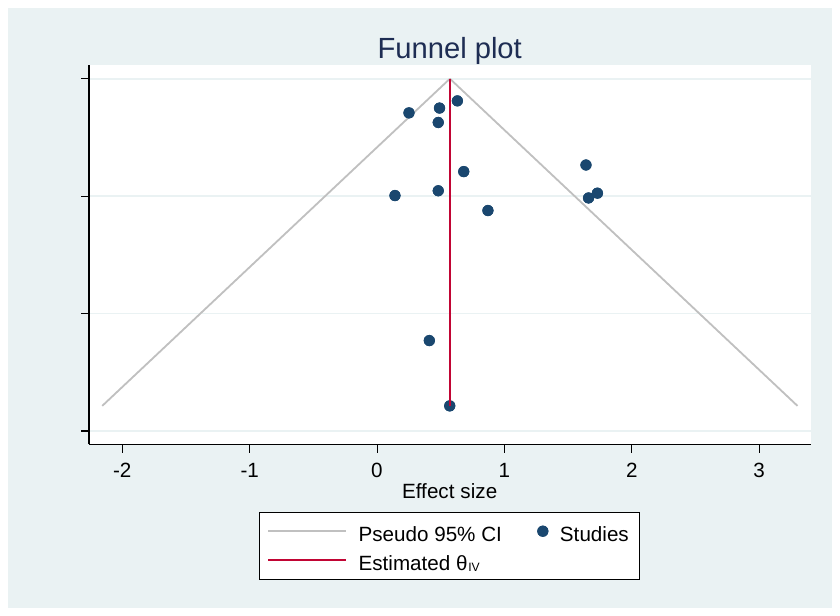

**eFigure 13. Funnel plot assessing publication bias in the association of mortality of COVID-19 with use of RAS inhibitors**

Unadjusted OR model (P_Egger‘s test_=0.19)

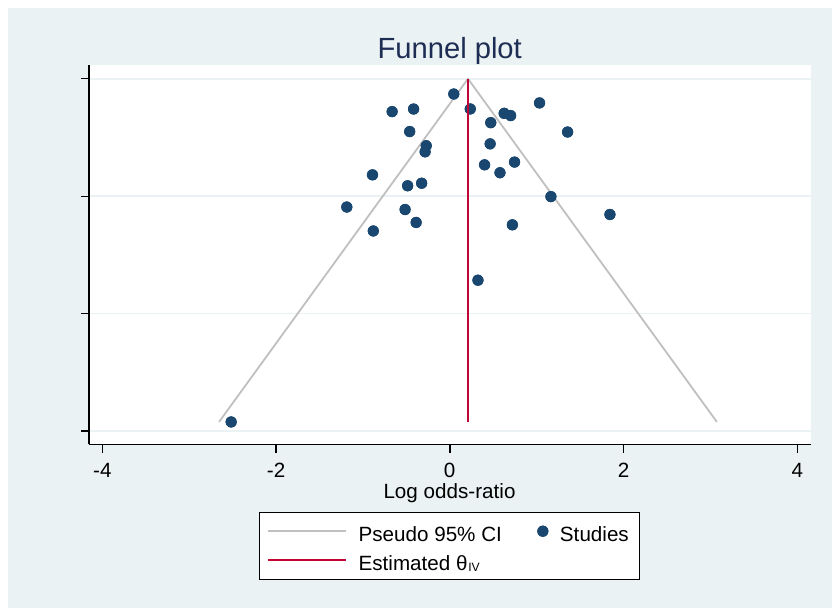

Adjusted OR model (P_Egger‘s test_=0.90)

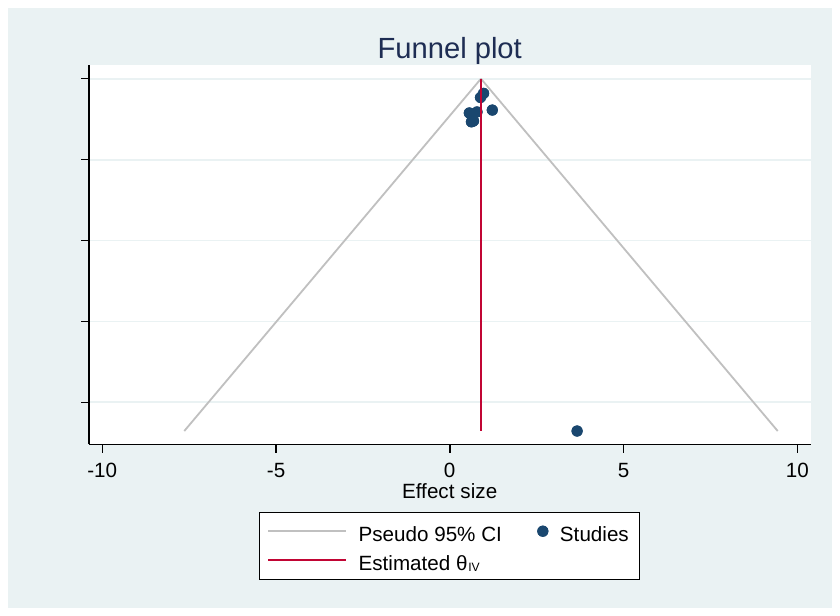

Adjusted HR model (P_Egger‘s test_=0.77)

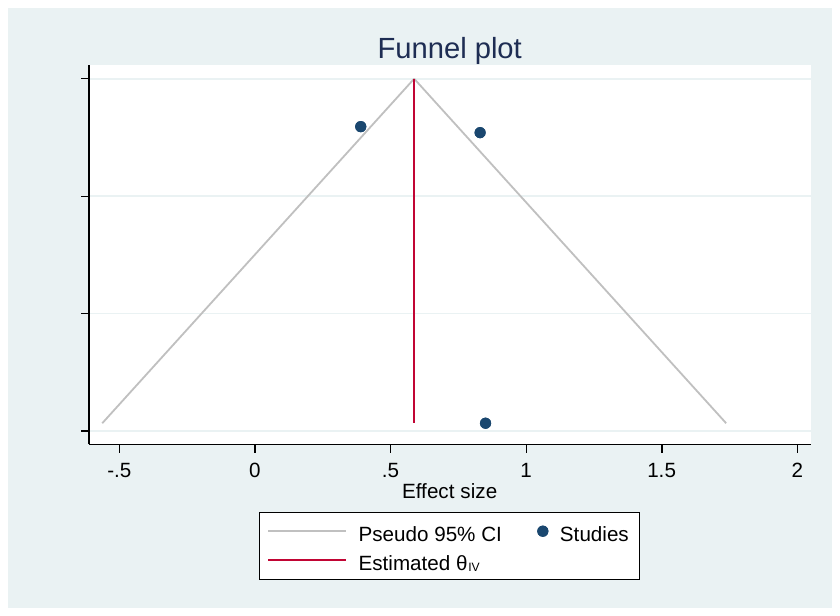
